# Impact of opinion dynamics on the public health damage inflicted by COVID-19 in the presence of societal heterogeneities

**DOI:** 10.1101/2023.03.26.23287758

**Authors:** Rex N. Ali, Saswati Sarkar

**Author notes:** Email addresses (Rex N. Ali), (Saswati Sarkar).

## Abstract

Certain behavioral practices such as wearing surgical masks, observing social distancing, and accepting vaccines impede the spread of COVID-19 and contain the severity of symptoms in the infected individuals. Opinions regarding whether to observe such behavioral practices evolve over time through interactions via networks that overlap with but are not identical to the physical interaction networks over which the disease progresses. This necessitates the joint study of COVID-19 evolution and opinion dynamics. We develop a mathematical model that can be easily adapted to a wide range of behavioral practices and captures in a computationally tractable manner the joint evolution of the disease and relevant opinions in populations of large sizes. Populations of large sizes are typically heterogeneous in that they comprise individuals of different age groups, genders, races, and underlying health conditions. Such groups have different propensities to imbibe severe forms of the disease, different physical contact, and social interaction patterns and rates. These lead to different disease and opinion dynamics in them. Our model is able to capture such diversities. Computations using our model reveal that opinion dynamics have a strong impact on fatality and hospitalization counts and the number of man-days lost due to symptoms both in the regular form of the disease and the extended forms, more commonly known as long COVID. We show that opinion dynamics in certain groups have a disproportionate impact on the overall public health attributes because they have high physical interaction rates, even when they have the lowest propensity to imbibe severe forms of the disease. This identifies a social vulnerability that mal-actors can utilize to inflict heavy public health damages through opinion campaigns targeting specific segments. Once such vulnerabilities are identified, which we accomplish, adequate precautions may be designed to enhance resilience to such targeted attacks and better protect public health.

## 1 Introduction

The novel coronavirus, COVID-19, was first detected in the United States in January 2020 [14]. Most of the detected COVID-19 in the early days were from travelers returning from high-risk countries or their close contacts. Thereafter, widespread community transmission of the disease began to occur in Washington, New York, and California communities [45]. On March 11, 2020, the World Health Organization (WHO) declared COVID-19 a pandemic [12]. Since then, there have been approximately 97 million cases and 1.06 million fatalities in the U.S. [2].

Behavioral patterns heavily influence both the spread and evolution of COVID-19. For example, a large-scale randomized trial involving 350000 people in 600 villages in Bangladesh has shown that the use of surgical masks impedes the spread of COVID-19 [19]. Observing isolation when exposed or infected also impedes the spread. Vaccines reduce the severity of symptoms, including hospitalization and death, and also spread rates [44]. Opinions regarding whether to observe behavioral patterns conducive to containment of COVID-19 evolve over time through social exchanges via networks that overlap with but are not identical to the COVID-19 propagation networks. Thus, the biological and information contagion spread simultaneously and necessitate a joint investigation of the two phenomena. The joint investigation is yet to be studied for COVID-19, despite enormous progress in research on COVID-19 in the last 3 years. To the best of our knowledge, our recent work on modeling the joint spread of Smallpox and vaccine hesitancy constitutes the only study on the joint spread of infectious disease and opinion pertaining to behavioral patterns that affect the spread of the disease [20]. But certain distinguishing characteristics of COVID-19 necessitate an investigation of the joint spread focusing on COVID-19.

Several behavioral patterns, such as willingness to wear surgical masks, observe social distance, affect the spread of COVID-19. Next, whether an individual is vaccinated affects the severity of symptoms in infected individuals [18, 42]. Next, the severity of COVID-19 symptoms and fatality rates depend on age [8, 16, 25], underlying health conditions (obesity, cancer, heart conditions, chronic liver disease, chronic lung disease, chronic kidney disease, HIV infection, etc.) [7, 26], race [29], gender of individuals [33], while propensity to spread COVID-19 depends on the contact patterns of individuals. More specifically, old individuals and those with certain underlying diseases, hitherto referred to as comorbidity are at greater risk to develop a severe form of the disease and even die from it, the risk factor is higher for males and for certain ethnic groups. But old individuals are also least likely to spread the disease because their contact rates are limited [37]. Young individuals who are in good health usually have high contact rates among themselves, and also with other groups, and are therefore more likely to spread the disease. But they are also least likely to develop a severe form of the disease. Given this divergence between groups who are at greater risk from the disease and who are most likely to spread it, there may also be a divergence between opinions regarding behavioral patterns between them. That is, given that their risk factors for developing a severe form of the disease are lower, young and healthy individuals may more easily subscribe to opinions that undervalue behavioral patterns that are conducive towards containment of the disease. Opinion spread rates can be different in different groups too. For example, young individuals may be more active on social media and may therefore circulate their opinions faster than others. The groups are often clustered in that the contact rates (both proximity and social contacts) are often higher within groups than across groups. For example, residential neighborhoods and social circles are often racially segregated [43] - according to a poll conducted in 2013 by Reuters/Ipsos, about 40 percent of white Americans and about 25 percent of non-white Americans are surrounded exclusively by friends of their own race [30]. Thus, the joint disease and opinion spread need to be studied considering different forms of behavioral patterns that affect the spread and different types and rates of information and disease spread based on age, underlying health conditions, gender, race, etc. Note that the divergences in question need not be as pronounced for other infectious diseases as COVID-19.

The emergence of a new infectious disease always opens up the possibility of a new bioterrorist attack that utilizes the respective contagion. It is possible to create synthetic strains of COVID-19 that are more virulent and has higher transmissibility than the existing strains through gain-of-function (GOF) research. One such strain has already been created by a group of researchers at Boston University (BU) by combining two features of different existing COVID-19 strains [27]. GOF research may help researchers better understand the evolving pathogenic landscape, develop new technologies, pandemic response, treatment, and countermeasures, but may lead to synthetic variants which can be leaked accidentally or deliberately as part of a bioterrorist attack. In the event of the latter, opinions may be manipulated to induce behaviors conducive to the spread of the disease. The divergence between groups that are at the greatest risk and can spread the most facilitates such behavior manipulation through opinion spread. For example, simultaneously with the deliberate seeding of synthetic strains in the populace, behavior conducive to the spread may be promoted among young individuals citing their low risk of the disease. Due to connection across age groups, the disease may spread to old and at-risk individuals leading to high casualty. Thus, the characteristics of COVID-19 may be exploited to launch lethal attacks through the spread of opinions along with the spread of disease. The potential of malevolent actors to inflict damage by exploiting the above characteristics needs to be investigated.

In this paper, we develop a computationally tractable mathematical model that jointly captures the evolution of a COVID-19 outbreak and the evolution of opinion pertaining to behavioral patterns pertinent to the spread and severity of the disease. The model captures 1) the distinguishing characteristics of COVID-19, namely different risk factors and contact patterns of different sections of the populace classified through age, underlying health conditions (comorbidity), race, and gender; 2) different opinion dynamics and rates of spreads of opinions within these groups and across the groups. The model is flexible enough to capture the dynamics of different kinds of behavior, namely wearing surgical masks, and receiving vaccines. We consider the reality that in the age of social media, opinions regarding behavioral dynamics rapidly evolve, through social networks that overlap with but are not identical to biological networks. In particular, during physical interactions, both disease and opinions may spread, whereas only opinions may spread through remote (e.g., electronic) interactions, and only the disease might spread when individuals share the same physical space (e.g., public spaces like beaches, parks, public transports) without engaging in social interactions. We consider the long-term effects of COVID-19 infection, otherwise known as long COVID or post-COVID conditions (PCC). As per WHO, long COVID occurs when coronavirus symptoms persist or return three months after an individual develops symptoms, and the symptoms last for at least 2 months and cannot be explained by an alternative diagnosis [17]. Common symptoms of PCC include fatigue, shortness of breath, and cognitive dysfunction. According to the CDC, unvaccinated individuals who get infected with COVID-19 might be at higher risk of developing long COVID compared to people who get infected after receiving the vaccine [6]. Thus, PCC is correlated to behavioral choice and therefore opinion dynamics. The model captures the essence of stochastic evolution, but retains computational tractability, in that it easily scales to typical target population sizes for infectious diseases encompassing millions of individuals. We utilize the model to quantify the impact of the opinion dynamics on metrics that capture the overall health of the system such as the total number of fatalities, hospitalization, and the number of days the populace suffers from symptoms. This provides a quantitative foundation for public health discourse pertaining to the relationship between disease and opinion spreads that have only been conducted in the qualitative sphere for COVID-19 thus far. The quantifications confirm several common-place intuitions and go beyond by unearthing the exact nature of the dependence of the above public health metrics on several key parameters. Our work helps identify how potential bioterrorist attacks can strategically exploit the specific characteristics of COVID-19 to undermine public health.

## 2 Materials and Methods

In this Section, we progressively describe the dynamics of COVID-19 progression and spread (Section 2.1), the spread of opinions pertaining to different behavioral patterns that affect the severity of symptoms and the rate of spread (Section 2.2), state transitions in the joint spread of disease and opinions (Section 2.3), a mathematical formulation that captures the dynamics of the joint spread (Section 2.4).

### 2.1 Dynamics of the infectious disease

SARS-CoV-2 propagates through spatial proximity. When a *susceptible* individual, *S*, gets in spatial proximity of an *infected* individual, *I*, the former contracts the disease with a certain probability, and becomes *exposed*. We denote the exposed stage by *E*. The virus incubates and grows during the exposed stage but the exposed individual neither shows any symptoms nor is contagious. From the exposed stage an individual either becomes *presymptomatic*, stage denoted by *P* , or *asymptomatic*, stage denoted by *I_a_*. During both stages, the individual does not show symptoms but is infectious, though he may transmit the disease with different probabilities. From the former stage, he moves on to show symptoms while from the latter stage, he directly recovers without ever showing symptoms. If he shows symptoms, he transitions into the *symptomatic* stage *I_s_* after which he either recovers or is hospitalized. When he is hospitalized, he can not transmit the infection further because he is under quarantine. A hospitalized individual, stage *H*, can recover, stage *R*, or die, stage *D*. We assumed that an individual who recovers from COVID-19 does not get re-infected. Refer to Fig 1 for a pictorial illustration of the different stages of the disease.

**Fig 1.**
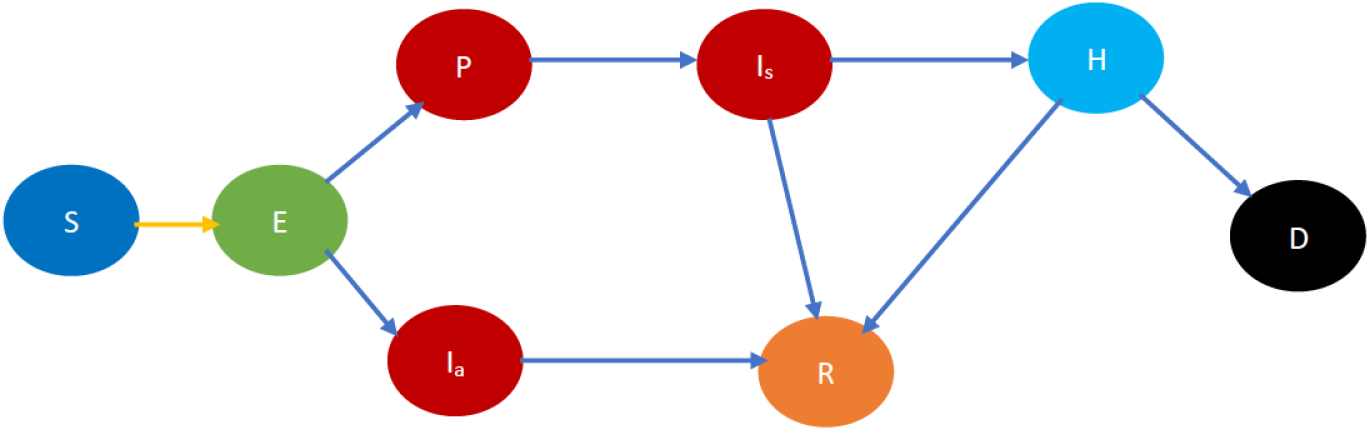
Fundamental COVID-19 state diagram. The states in blue color are the susceptibles (not yet infected but they are prone to infection) while those in light green color are exposed (still in the incubation period, hence they are not infectious). The states in dark red are infected and therefore infectious while those in gold have recovered, those in light blue are hospitalized, and black denotes dead. In addition, the yellow arrows show susceptibles transitioning to the exposed state after contracting the virus. The blue arrows indicate the natural progression of the disease.

One of the distinguishing characteristics of COVID-19 is that different segments of the populace exhibit a substantially different propensity to severe forms of COVID-19, and death when they imbibe it. First, once infected, older adults are more likely to need hospitalization, intensive care admission, or a ventilator to help them breathe [16]. Older patients also have a higher fatality rate [8]. Next, male patients are more at risk for worse outcomes and death [33]. Also, in the U.S. once infected, African Americans have higher duration of hospital stays, as compared to white people [29]. Finally, risks for a severe form of the disease are substantially higher when patients have certain underlying health conditions. These underlying conditions include obesity (Body Mass Index, i.e., a person’s weight in kilograms divided by the square of height in meters, over 30 is considered obese [4]), cancer treatment, HIV/AIDS positive, heart and lung disease, diabetes, etc [26]. Risks increase manifold for comorbid patients, i.e., those who suffer from two or more conditions.

To model different risk factors for different sections of the population, we will classify individuals in different stages of COVID-19 further according to their age, gender, race, and underlying health conditions. Specifically, we classify individuals into three different age groups: (i) youngest group - for individuals who are in the age range of 0 *−* 24 years; (ii) middle age group - for individuals who are 25 *−* 49 years old; (iii) oldest group - for those who are 50 years and above. We classify individuals into male and female, African-American, Hispanic and white, healthy and immunocompromised. Individuals who do not have any underlying health conditions that accentuate the risk of imbibing a severe form of COVID-19 are classified as healthy, the rest are immunocompromised.

Refer to Fig 2 for a pictorial illustration of the evolution when only age-based classification is resorted to. Note how the state evolution shown in Fig 1 is being replicated due to age-based classification. Consideration of further classification based on gender, race, and underlying health replicates the state evolution shown in Fig 2 further. We consider this further classification in the mathematical formulation but not in the pictorial representation owing to the large number of states introduced due to the full-fledged classification.

**Fig 2.**
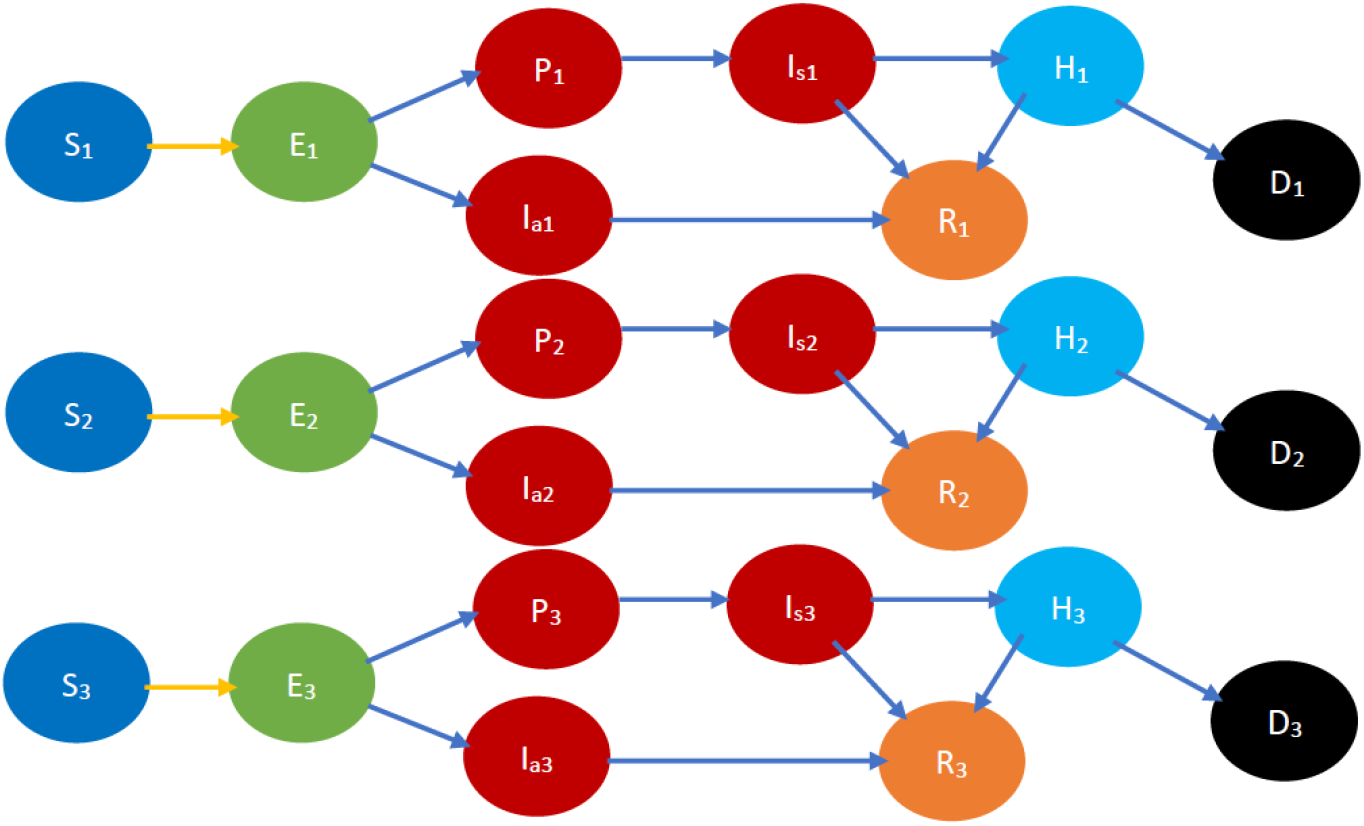
COVID-19 state diagram divided into various age groups. The subscripts 1, 2, and 3 represent the youngest group, middle age group, and oldest group respectively. Every state and transition has the same meaning as described in the caption for Fig 1. Note that there is no transition between states indexed 1, 2, 3, e.g., between *S*_1_*, S*_2_*, S*_3_ as we don’t consider transitions between age groups in the time frame under consideration.

Different sections exhibit different contact rates and patterns, both within and across sections. For example, in many parts of the United States, neighborhoods are racially segregated, through living preferences and economic conditions. Accordingly, people of different races also largely attend different schools, and utilize different services and shops [43]. According to a poll conducted in 2013 by Reuters/Ipsos, about 40 percent of white Americans and about 25 percent of non-white Americans befriend exclusively from their own race [30]. Thus, contact rates across the categories we considered in the previous paragraph are lower than those within the same category. Finally, contact rates also depend on age. Specifically, older people have less social contact than younger people, and other things being equal, higher age is correlated with a lesser amount of time spent with others [37]. Different contact rates and patterns can be incorporated by classifying individuals based both on the stage of the disease and age, gender, race, overall health, etc.

### 2.2 Opinion dynamics

We now describe how we characterize the opinion dynamics leading to a characterization of the joint evolution of COVID-19 and opinions.

An individual is *cooperative* if he follows the behavioral practices that reduce 1) the probability of the spread of COVID-19 in the event of spatial proximity between an infectious and a susceptible individual; 2) the severity of symptoms if he imbibes the disease. Such behavioral practices include some or all of the following: maintaining appropriate physical distancing, wearing appropriate protective gear, getting vaccinated, frequent hand washing, etc. A *non-cooperative* individual does not observe these norms. Thus, an individual ought to be classified as cooperative or non-cooperative, in addition to his stage of the disease, age, gender, race, and overall health. For example, in the classified COVID-19 state diagram (Fig 3), *S_c_* and *S_n_* respectively denote cooperative and non-cooperative susceptible individuals. This additional classification enables the modeling of the joint evolution of the disease and opinions. Refer to Supporting Information Table 1 for all the notations.

**Fig 3.**
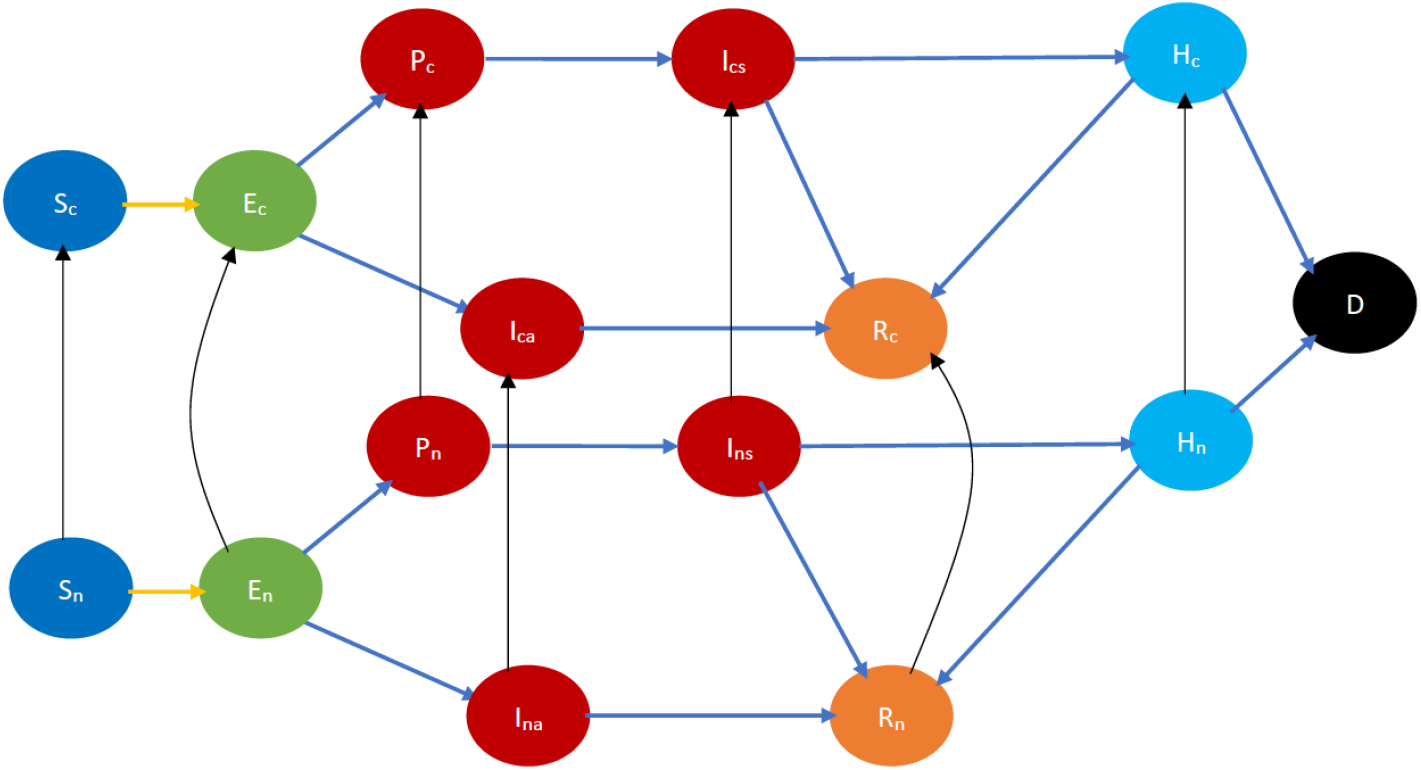
COVID-19 state diagram incorporating cooperativity. The black arrows indicate opinion evolution. The subscript *c* and *n* respectively denote cooperative and non-cooperative individuals. Thus, the black arrow from *S_n_* to *S_c_* represents the conversion of a susceptible non-cooperative to a susceptible cooperative, i.e., evolution of opinion of a susceptible. Every state and transition has the same meaning as described in the caption for Fig 1.

**Table 1.** Fundamental compartments of the model

| Compartment | Description |
| --- | --- |
| $S_c$ | Cooperative susceptible |
| $S_n$ | Non-cooperative susceptible |
| $E_c$ | Cooperative exposed individuals |
| $E_n$ | Non-cooperative exposed individuals |
| $P_c$ | Cooperative presymptomatic individuals |
| $P_n$ | Non-cooperative presymptomatic individuals |
| $I_{cs}$ | Cooperative symptomatic infected individuals |
| $I_{ns}$ | Non-cooperative symptomatic infected individuals |
| $I_{ca}$ | Cooperative asymptomatic infected individuals |
| $I_{na}$ | Non-cooperative asymptomatic infected individuals |
| $H_c$ | Cooperative hospitalized individuals |
| $H_n$ | Non-cooperative hospitalized individuals |
| $R_c$ | Cooperative recovered individuals |
| $R_n$ | Non-cooperative recovered individuals |
| $D$ | Dead |

Opinions pertaining to the observance of the above behavioral practices evolve through the influence of social contacts and through exposure to independently accessible information such as reading material, TV, Radio, billboards, internet advertisements, etc. We consider that a cooperative converts a non-cooperative to become a cooperative with a certain probability during an exchange of ideas between the duo. Such exchanges may happen in person or remotely. A non-cooperative may also be converted by reading about what reduces the spread of the disease or by watching or hearing public awareness programs on TV, radio, billboards, social media, etc. The conversion may happen during any stage of the disease. We assume that when an individual is hospitalized, he is quarantined, then he does not convert anyone or is converted by anyone but may be converted by exposure to public awareness campaigns (which may have a stronger impact because the individual might be infected and he experiences the symptoms acutely). Our default assumption is that the non-cooperatives change their opinions, but we later generalize the model to incorporate change of opinions in the reverse direction as well.

Refer to Fig 3 for a pictorial illustration of the evolution when only cooperativity-based classification is resorted to. Note how the state evolution shown in Fig 1 is being replicated due to the cooperativity-based classification. Fig 4 shows classification based on both age and cooperativity. Further classification based on age, gender, race, and underlying health, which we consider in the mathematical formulation, replicates the state evolution shown in Fig 3 further. The fundamental difference between classifications based on cooperativity and those based on age, gender, race, and underlying health is that an individual can transition from cooperative to non-cooperative or vice-versa but does not transition across the other attributes in the time frame we consider. Thus, in Fig 2 there is no transition between *S*_1_*, S*_2_*, S*_3_, but in Fig 3 there is a transition from *S_n_* to *S_c_*.

**Fig 4.**
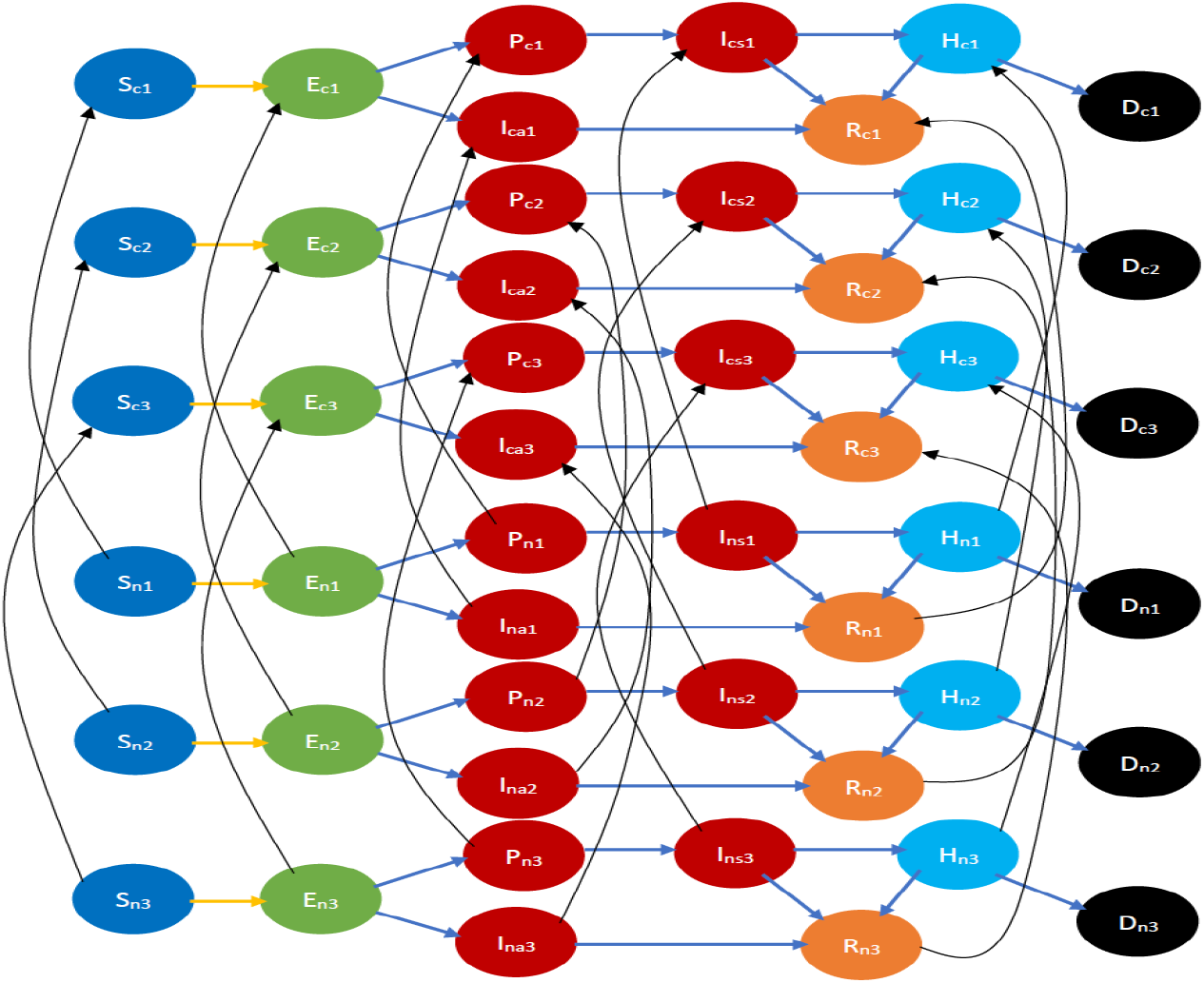
COVID-19 state diagram classified by ages and cooperativities. The subscript *c* and *n* respectively denote cooperative and non-cooperative individuals. Similarly, subscripts 1, 2, and 3 represent the youngest group, the middle age group, and the oldest group respectively. Note that there is no transition across age groups, but there is a transition across opinion groups, e.g., there is a transition from *S_n_*_1_ to *S_c_*_1_, but no transition between *S_n_*_1_*, S_n_*_2_, or *S_c_*_1_*, S_c_*_2_. Every state and transition has the same meaning as described in the caption for Fig 1.

We have considered different behavioral practices pertaining to the evolution of COVID-19, namely: maintaining appropriate physical distancing, wearing appropriate protective gear, frequent hand washing, getting vaccinated, etc. All except the last reduce the probability of spread of the disease from an infectious individual to a susceptible, while the last reduces the severity of symptoms, probability of hospitalization, and death once the disease is imbibed (though the last may also reduce the probability of transmission of the disease from the infectious to the susceptible). We now illustrate how minor adaptations of the model can help cater to these distinctions in the impact of different behavioral precautions. We first consider the behavioral practices that only reduce the spread. When cooperativity is used to denote observance of these, we assume that when an infectious individual is in physical proximity of a susceptible one, the probability of the spread of the disease from the infectious to the susceptible is higher if both are non-cooperatives than if both are cooperatives. If only one is non-cooperative, the probability of the spread is somewhere in between (the in-between can also equal one of the two ends, i.e., the probability when both are cooperative and when both are non-cooperative). We now consider the behavioral practice to be vaccinated, i.e., a cooperative is willing to be vaccinated while a non-cooperative is not. We assume that vaccines are available as soon as one is willing, which is currently the case in most of the US and Europe. Thus, considering that the vaccine in question requires only one dose, as soon as an individual becomes cooperative, he is vaccinated. Then, we incorporate the impact of vaccination by considering that a cooperative has a lower probability of developing symptoms, and a lower probability of needing hospitalization even when he develops symptoms (i.e., the symptoms are mostly mild), and a lower probability of dying even when hospitalized. We can simultaneously consider that cooperatives spread the disease with a lower probability, as described above if vaccines also lower the probability of the spread. Our default assumption is that the vaccine needs only one dose, though we describe in Section 4 how our model can be generalized to incorporate vaccines that need multiple doses. The model can incorporate different opinion dynamics and interactions involving exchange of ideas in different sections of the populace based on age, gender, race, health conditions, by considering different rates of conversion of opinions in different groups representing the above classifications.

### 2.3 State transitions in the joint evolution of disease and opinions

There are two kinds of transitions in joint evolution: (1) interactional and (2) non-interactional. Interactional transitions happen when a susceptible imbibes the disease from an infectious individual or a cooperative converts a non-cooperative. Thus, during interactional transition, both disease and opinions may spread from one individual to another. The natural progression of the disease in the infected individuals and conversion of opinion due to access to information in reading material or mass awareness campaigns constitute non-interactional transitions.

We now provide a taxonomy of the interactional transitions (which is similar to that in [20]). Interaction between individuals can be classified as (i) physical interactions with an exchange of opinion and biological contagion (e.g., friends and acquaintances visiting homes of each other, etc.); (ii) physical interactions without any exchange of ideas (e.g., people commuting on a train, bus, etc.); (iii) virtual interactions with an exchange of ideas (e.g., telehealth, counseling over the phone or internet, etc.). Whereas the first case can cause infection and a change in opinion, the second case can only cause infection, while the third case can only cause a change of opinion.

### 2.4 The Clustered Epidemiological Differential Equation (CEDE) model

A joint investigation of infectious disease and opinion dynamics naturally leads to a computationally complex model involving 1) a multiplicity of states representing a combination of stages of the disease, gender, age group, race, health condition, and cooperativity; and 2) a multiplicity of state transitions representing interactional transitions due to spread of the disease and opinions, as well as non-interactional transitions due to the natural progression of the disease in the infected individuals and alteration of opinions due to exposure to public awareness campaigns. Following our recent work on modeling the joint spread of Smallpox and vaccine hesitancy [20], we model these attributes by adapting the *metapopulation epidemiological model* [22,28] which relies on a set of differential equations. We describe the adaptations in the Supporting Information and refer to the resulting model as *clustered epidemiological differential equations* or the CEDE, given that each state is decomposed into groups or clusters. In general, estimating the spread of infectious diseases is computationally challenging because it involves millions of individuals in population sizes one needs to consider. Metapopulation models alleviate this challenge by relying on differential equations, which can be solved using readily available and computationally efficient numerical techniques. The differential equations capture the evolution of states of different fractions of the total population. Thus, the computational time does not increase with the increase in the size of the populace [20].

Each variable in the CEDE represents the fraction of the population who are in a particular system state. Meanwhile, each state represents the combination of the stage of the disease, age group, gender, health conditions, and cooperativity of an individual. Each differential equation captures the evolution of a particular variable. Thus, the solution of the CEDE provides the fraction of individuals in different states at given times. In other words, the solutions of the differential equation provide the distribution of the disease and opinion spread across groups and time. The terms in the CEDE are either linear or quadratic. The quadratic terms represent the interactional transitions (refer to the yellow and black arrows in Fig 3) and the linear terms represent the non-interactional transitions (refer to the blue arrows in Fig 3). This distinction arises because interactions always involve two individuals, while non-interactional transitions involve only one individual. This arises in all epidemiological models starting from the classical Kermack–McKendrick formulation [34] and onward to the metapopulation models [20, 22, 28, 31, 35]. But our work differs from the metapopulation epidemiological models because it captures two different evolving contact processes spreading simultaneously (disease, opinion), whose overlap only partially (only certain interactions spread both the disease and opinion, while others can only spread one of the two). The spread of the two processes involves two broad categories of interactional transitions: (1) susceptible to exposed state; (2) non-cooperative to cooperative and vice versa. Metapopulation models typically capture only one evolving process, namely the disease, spreading through physical proximity and therefore need only the first kind of interactional transitions. Since interactional transitions are represented by quadratic terms, the second kind produces additional quadratic terms in our model.

We now provide more details on the computation time of the CEDE. Let there be *L* variables and thus *L* differential equations. The computation time depends only on *L* and increases linearly with it. For instance, with 3 age groups, 2 genders, 3 races, cooperative-non-cooperative, healthy-immunocompromised classifications, *L* = 450. And, regardless of the population size, the average computation time to obtain one data point is approximately 2 minutes using MATLAB 2022a and Macbook Pro 2.9 GHz Dual-Core Intel Core i5 Processor with 8 GB Memory Laptop.

A question that arises for the CEDE is that it is a deterministic model, while many of the state transitions are stochastic. We resolved this dilemma in our earlier work [20] through an application of a classical result of probability theory. Under some commonly made assumptions on the stochastic evolutions, we have shown that as the number of individuals increases, the fractions of individuals in different system states in the stochastic system converge to the solutions of the CEDE, and the convergence becomes exact in the limit that the number of individuals is infinity [20]. Thus the CEDE approximates the stochastic process better with an increase in the number of individuals. The convergence guarantee holds when the stochastic evolutions are *Markov*, that is, the amount of time an individual spends in each system state is *exponentially* distributed, which is what we assume to estimate the parameters of the system (Supporting Information). Note that such Markovian assumptions are commonplace in modeling the spread of infectious diseases (e.g., as noted in Chapter 2, p. 28, [32]).

Our CEDE model is naturally flexible in that it can accommodate opinion dynamics, different behavioral practices, arbitrary age groups, gender, health conditions, and countermeasure application strategies. The CEDE remains computationally tractable and provides an analytical convergence guarantee while providing the above modeling flexibility.

## 3 Results

We utilized our COVID-19 model to investigate the impact of opinion dynamics on various public health attributes such as fatalities, number of man-days lost because of symptoms, hospitalizations, etc. We consider three different scenarios: 1) cooperatives observe behavioral practices (e.g., wearing surgical masks, observing social distancing, hygiene such as repeated hand washing) that reduce the spread of the biological contagion (Scenario 1, Section 3.2); 2) cooperatives take vaccines which reduce the severity of symptoms (Scenario 2, Section 3.3); 3) cooperatives observe both healthy behavioral practices and take vaccines (Scenario 3, Section 3.4). Recall that consistent with reported statistics, we have assumed that vaccines only reduce the severity of symptoms, thus in Scenario 1 cooperatives only impede the spread while in Scenario 2 they only have a lower propensity of severe symptoms, in Scenario 3 they impede the spread, and have a lower propensity of severe symptoms [18, 19, 42, 44]. In the first and third scenarios, our default assumption is that cooperative susceptibles (respectively cooperative infectious) individuals are infected (respectively infect) during physical proximity with an infectious (respectively susceptible) individual at a rate that is half that of a non-cooperative individual. We also deviate from the default assumption and consider the scenario in which when a susceptible is in physical proximity of an infectious and 1) both are cooperatives, the disease transmits at the lowest rate; 2) only one is a cooperative, the disease transmits at an intermediate rate; and 3) both are non-cooperatives, the disease transmits at the highest rate. We consider that the lowest and the intermediate are respectively 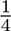 and 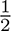 of the highest rate. We denote this scenario as *second-transmission-rate-scenario*. We consider different durations for COVID-19 symptoms: 1) regular durations in which symptoms do not exist after an individual recovers; 2) long COVID in which symptoms last for months after recovery (we consider two months in this case). We consider different fatality and hospitalization rates, ranging from those seen in natural variants (default choice) to extremely high values which have been reported for synthetic variants. As to information dynamics, we consider that cooperatives convert non-cooperatives during information exchange (default), but also we consider the scenario in that non-cooperatives convert cooperatives.

We start with a sanity check in which we show that the several commonly used variables such as the number of infected, hospitalized, and recovered individuals vary with time in an expected manner (Section 3.1). This suggests that the model is functioning as per expectations and instills confidence in the results that the model yields. We subsequently report our findings for the above-mentioned variations in Sections 3.2, 3.3, and 3.4. In all the variations, we observe that information exchange between cooperatives and non-cooperatives have significant effect on the public health attributes we consider. Specifically, information exchange among the younger population has a disproportionate impact on public health attributes, though statistically speaking the symptoms are the mildest among them. The disproportionate impact arises because the younger section is in physical proximity to each other and with the rest of society more often than other age groups, because of their naturally active lifestyle. Unless otherwise mentioned, all parameters are at their default values specified in Supporting Information Section II. Figs 16 - 20 are supplementary figures in the Supporting Information.

### 3.1 Sanity check

We plot the number of infected, hospitalized, and recovered individuals as a function of time (Scenario 1, Fig 16, Scenario 2, Fig 17, Scenario 3, Fig 5). We plot these attributes separately for the youngest and the oldest age groups we considered. As expected, the first two increase with time initially and then decrease, as individuals either recover or die. The number of recovered individuals initially increases with time and finally saturates as the system reaches a steady state in which all individuals have either recovered or died. Finally, as expected, more individuals in the youngest group are infected per day than individuals in the oldest group (Figs 16a, 17a, 5a). This is because younger individuals are more often in physical proximity with others than those in the oldest group. As expected, the number of hospitalized individuals is higher for the oldest group because the hospitalization rate increases with age (Figs 16b, 17b, 5b). The number of recovered individuals is higher for the youngest group because the fatality rate is lowest for them and in the steady state an individual is either dead or recovered (Figs 16c, 17c, 5c). The number of infected and hospitalized are the least and the number of recovered the maximum in Scenario 3, intermediate in Scenario 1, and respectively the maximum and the minimum in Scenario 2 (Figs 16, 17, 5). This is because the cooperatives observe all the desirable behavioral practices in Scenario 3 and different subsets of behavioral practices in the other two Scenarios. A comparison between Scenarios 1 and 2 suggests that impeding the spread is more effective than reducing the severity of symptoms for all public health attributes (Figs 16, 17).

**Fig 5.**
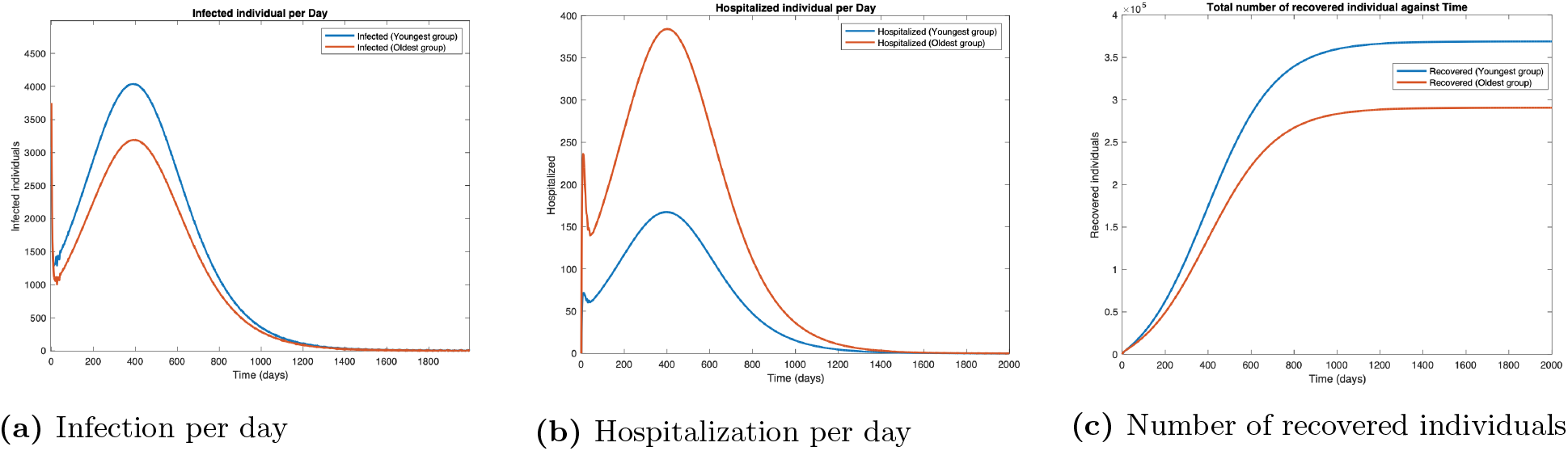
Scenario 3. Plots in the time domain for sanity check.

### 3.2 Impact of opinion dynamics when behavioral practices affect spread of COVID-19: Scenario 1

#### 3.2.1 The default scenario

We consider that cooperatives observe behavioral practices that reduce the spread of biological contagion, while non-cooperatives do not observe such practices. No individual takes vaccines. In Fig 6 we plot the overall fatality and hospitalization counts against the opinion spread rate for different values of the fraction of individuals who are cooperative initially. As cooperatives exchange opinions with non-cooperatives, the latter are converted. In general, the conversion happens faster as the rate of exchange of opinions, i.e., the opinion spread rate, increases. The conversion does not happen if all individuals are cooperatives or non-cooperatives initially. Thus the number of deaths does not change with the opinion spread rate when the initial cooperativity is either 0 or 1. The fatality counts however decrease rapidly with an increase in opinion spread rates for initial cooperativities between 0 and 1. For instance, for an initial cooperativity of 0.8, 0.6, 0.4, and 0.2, the number of deaths decreases by 42.33%, 95.06%, 97.35%, and 97.36% respectively as the opinion spread rate increases from 0 through 2 *×* 10*^−^*^9^. Similarly, the number of hospitalized individuals decreases by 44.22%, 95.28%, 97.40%, and 97.37% respectively as the opinion spread rate increases from 0 through 2 *×* 10*^−^*^9^. The percentage decrease in fatality and hospitalization counts show that opinion dynamics have a strong impact on these public health attributes, the impact is strongest when the initial cooperativity values are moderate to low as then there are more non-cooperatives to convert with an increase in opinion exchange rates. The variations of the fatality and hospitalization counts against the opinion spread rate follow similar patterns.

**Fig 6.**
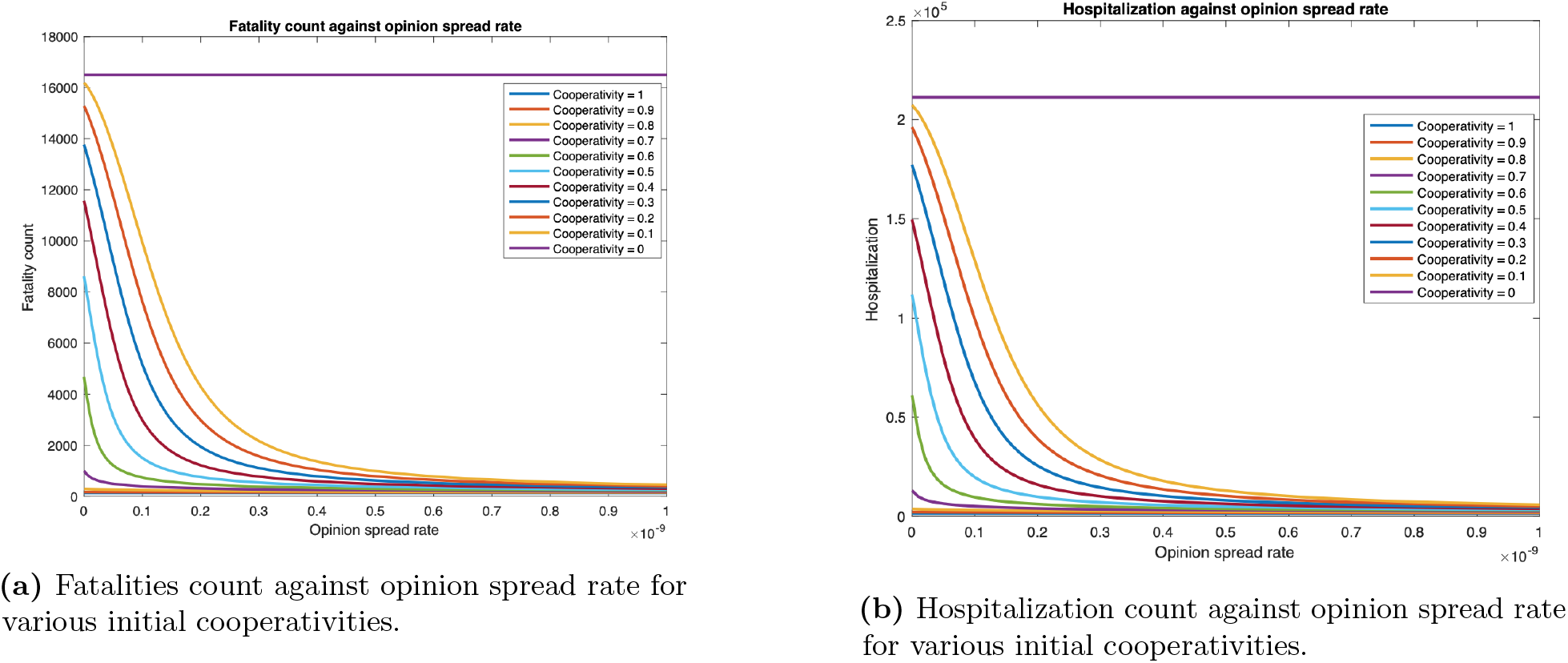
Fatality and hospitalization counts against opinion spread rate for various initial cooperativities.

We next examine how fatality and hospitalization counts are affected if only the opinion exchange rate in one age group is increased and also if there is any difference depending on which age group that is. We examine this for two different values of physical contact rates among the youngest population: the default value (*y_L_*) and thrice the default value (*y_H_* ). For *y_L_,* the fatality count in all, youngest, middle age, and oldest groups respectively decrease by 88.22%, 90.43%, 88.35%, and 88.10% as the opinion spread rate of the youngest group increases from 0 through 2 *×* 10*^−^*^9^ (Fig 7a). Also, the hospitalization count in all, youngest, middle age, and oldest groups respectively decreased by 88.66%, 90.30%, 88.37%, and 88.11% (Fig 18a). For *y_H_ ,* the number of deaths in all, youngest, middle age, and oldest groups will respectively decrease by 47.75%, 52.95%, 44.66%, and 47.63%. Similarly, hospitalization count in all, youngest, middle age, and oldest groups will respectively decrease by 48.60%, 52.87%, 44.70%, and 47.63%. Thus, an increase in the rate at which individuals physically interact reduces the percentage decrease in fatalities and hospitalization and thereby the efficacy of the opinion exchange. Note that the fatality and hospitalization counts for the oldest group are significantly affected by the opinion exchange within the youngest group. We now examine the impact of opinion exchange within the oldest group (Fig 7b). For *y_L_,* fatality count for all, youngest, middle age, and oldest groups decrease by 69.23%, 64.89%, 64.77%, and 69.80% respectively, as the opinion spread rate of the oldest group increases from 0 through 2 *×* 10*^−^*^9^. Also, the hospitalization count for all, youngest, middle age, and oldest groups will respectively decrease by 67.37%, 64.83%, 64.90%, and 69.81%. Next, for *y_H_ ,* the fatality count for all, youngest, middle age, and oldest groups will respectively decrease by 18.01%, 2.96%, 7.42%, and 19.92%. Similarly, the hospitalization count for all, youngest, middle age, and oldest groups will decrease by 11.75%, 2.94%, 7.43%, and 19.92% respectively. Thus, in all cases, the fatality and hospitalization counts decrease more when the opinion exchange rates are enhanced in the youngest group than when they are enhanced in the oldest group. Thus, converting younger individuals to cooperatives decreases fatality and hospitalization counts more across all age groups. That is, the behavioral practices of the youngest group impact the whole population more than those of the others. As expected, simultaneously converting opinions across all age groups have the most impact. For instance, for *y_L_* the fatality count for all, youngest, middle age, and oldest groups respectively decreased by 95.98%, 96.23% 96.59%, and 95.91% as the opinion spread rate increases from 0 through 2 *×* 10*^−^*^9^ (Fig 7c). Also, the hospitalization count for all, youngest, middle age, and oldest groups respectively decreased by 99.62%, 96.49% 93.66%, and 95.90%. Similarly, for *y_H_ ,* the number of deaths for all, youngest, middle age, and oldest groups decreases by 93.74%, 90.86% 93.62%, and 93.95% respectively. Also, the hospitalization count for all, youngest, middle age, and oldest groups respectively decrease by 92.91%, 90.85% 93.66%, and 93.95%. We do not present separate plots for overall fatality counts as the numbers can be obtained by adding the fatality counts in the three age groups; we just provide the numbers in the text. Yet again we observe from Figs 7 and 18 that the variation of hospitalization and fatality follows the same pattern. We, therefore, omit the plots for the latter subsequently.

**Fig 7.**
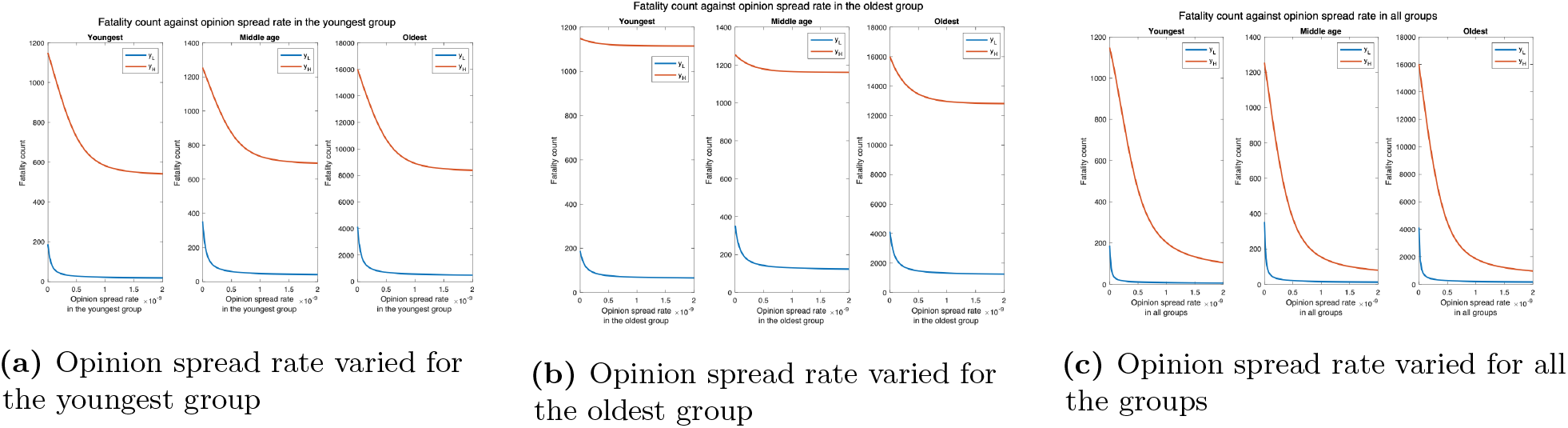
Fatality count for various interaction rates of the youngest group. *y_L_* denotes the default interaction rates of the the youngest group while *y_H_* represents when the youngest group’s interaction rates are tripled (other groups’ interaction rates remain the same).

To understand why the opinion exchange rate in the youngest group has the greatest impact on the fatality rates, we plot the fatality vs the opinion spread rate when the physical interaction rates are the same in the youngest and the oldest groups. This would clearly not arise in practice but has been selected so as to understand the reason underlying the phenomenon reported in the previous paragraph. We observe that the number of deaths for all, youngest, middle age, and oldest groups respectively decrease by 19.08%, 31.44%, 19.04%, and 18.74% as the opinion spread in the youngest group is increased from 0 to 10*^−^*^9^ (Fig 8a). Similarly, the number of hospitalized in all, youngest, middle age, and oldest groups respectively decrease by 38.14%, 27.98%, 28.15% and 38.95% as the opinion spread in the oldest group is increased in the same range (Fig 8b). The numbers are now in the same ballpark; if at all now fatalities decrease more by converting a greater number of cooperatives in the oldest group because death rates are higher here. Thus, in scenarios that arise in practice, the impact of the opinion spread in the youngest age group is the most because the physical interaction rate in this group is the highest, as such the disease spreads in this group at the highest rate and this group spreads the disease to other groups. Thus, converting individuals in this group to cooperatives in effect contains the carriers.

**Fig 8.**
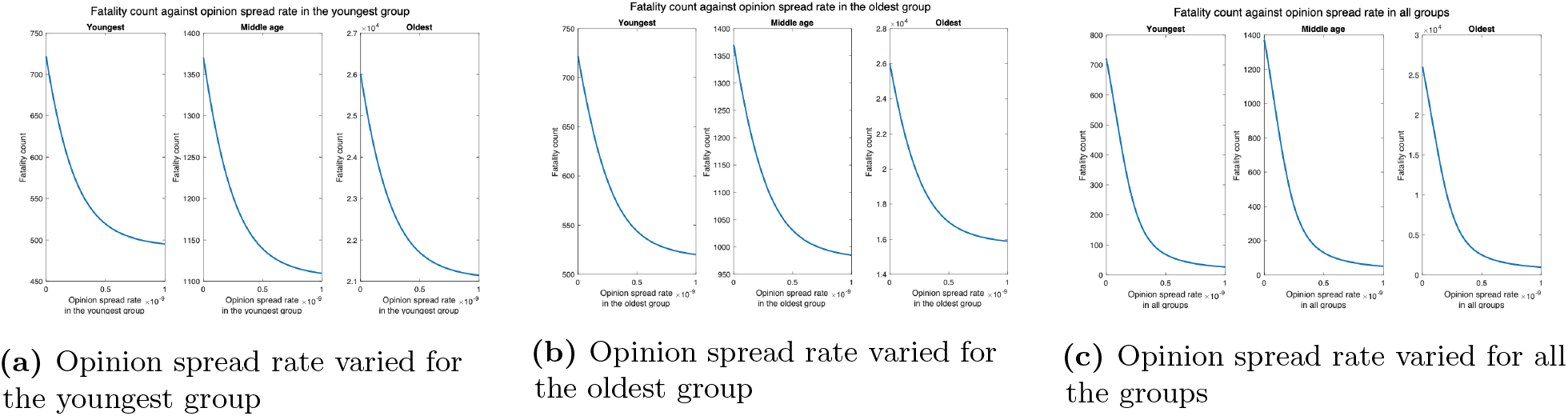
Fatality count when the interaction rates of the oldest group equal that of the youngest group.

#### 3.2.2 Second transmission rate scenario

Next, we consider the case when the disease spread rate is at its default value when only one of the interacting pair is cooperative; when both the interacting individuals are non-cooperative, the disease spread rate is twice the default value; and when both the interacting individuals are cooperative, the disease spread rate is half the default value. Fig 9a shows that the fatality counts for all, youngest, middle age, and oldest groups respectively decrease by 63.49%, 72.31%, 60.02% and 63.40% as the opinion spread rate of the youngest group increases from 0 through 2 *×* 10*^−^*^9^. Fig 9b shows that the fatality counts for all, youngest, middle age, and oldest group decrease by 46.36%, 28.12%, 28.36%, and 48.55% respectively, as the opinion spread rate of the oldest group increases from 0 through 2 *×* 10*^−^*^9^. Thus, the pattern of variation remains the same as before. Fig 9c shows that the fatality counts for all, youngest, middle age, and oldest groups respectively decrease by 98.81%, 98.85%, 98.85%, and 98.81% as the opinion spread rate for all the groups increases from 0 through 2 *×* 10*^−^*^9^. Finally even when the interaction rate of the youngest group is tripled, the fatality count for all, youngest, middle age, and oldest groups respectively decrease by 94.23%, 89.89, % 93.93%, and 94.51%. Thus, the impact of the opinion spread rate across all the groups is quite significant even when the disease spread rate is increased.

**Fig 9.**
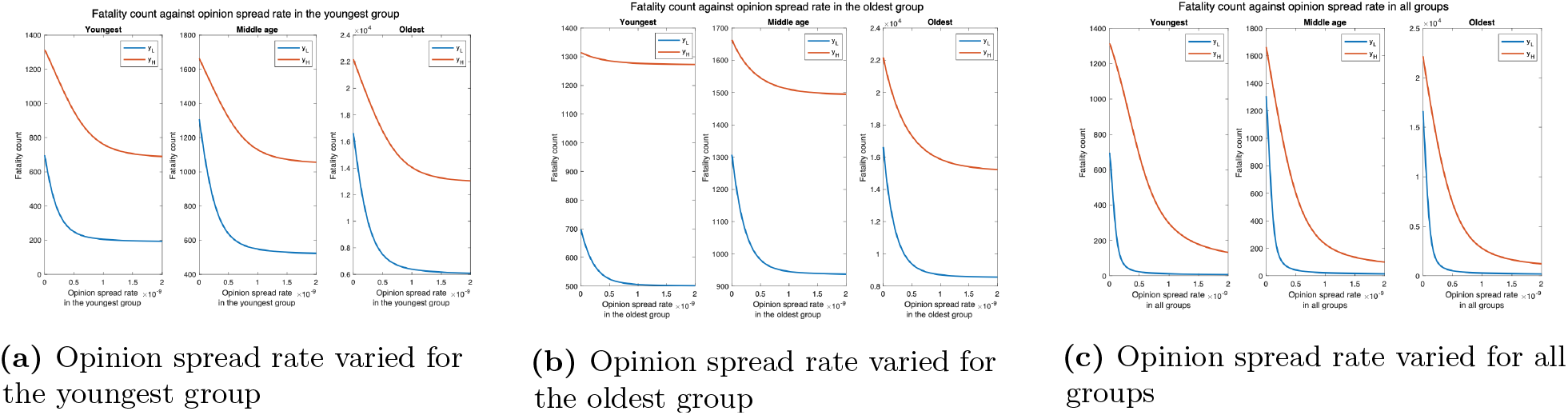
Fatality count for various interaction rates of the youngest group. *y_L_* denotes the default interaction rates of the the youngest group while *y_H_* represents when the youngest group’s interaction rates are tripled (other groups’ interaction rates remain the same).

#### 3.2.3 Impact of long COVID

Next, we evaluate the impact of opinion dynamics in presence of long COVID. Recall that long COVID occurs when coronavirus symptoms persist for at least 60 days or return three months after an individual becomes ill from SARS-CoV-2. We consider only the former here, in effect, this increases the duration of symptoms for patients or a fraction thereof and decreases the productivity of the populace. We consider the number of man-days patients suffer from symptoms, i.e., the number of days each patient suffers from symptoms summed up over all patients. This attribute is the sum of two components: 1) man-days lost due to symptoms during the symptomatic phase of COVID-19 infection; 2) man-days lost due to patients experiencing symptoms after recovering. Due to the second component, a section of the patients experiences symptoms even after transitioning to the recovered state when they are no longer infectious. The first component is 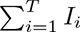, where *T* is the number of days we consider, *I_i_* is the number with symptoms, and the number hospitalized on day *i.* The second component is 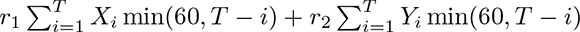, where *X_i_* is the number of cooperatives who recover on day *i*, *Y_i_* is the number of non-cooperatives who recover on day i, *r*_1_ is the fraction of cooperatives who suffer long COVID, *r*_2_ is the fraction of non-cooperatives who suffer long COVID. In the current section, neither cooperatives nor non-cooperatives are vaccinated. We consider *r*_1_ = *r*_2_ = 0.418 as 41.8% of unvaccinated individuals suffer from long COVID [23, 24]. We now plot the number of man days patients experience symptoms against the rate of spread of opinion (Fig 10).

**Fig 10.**
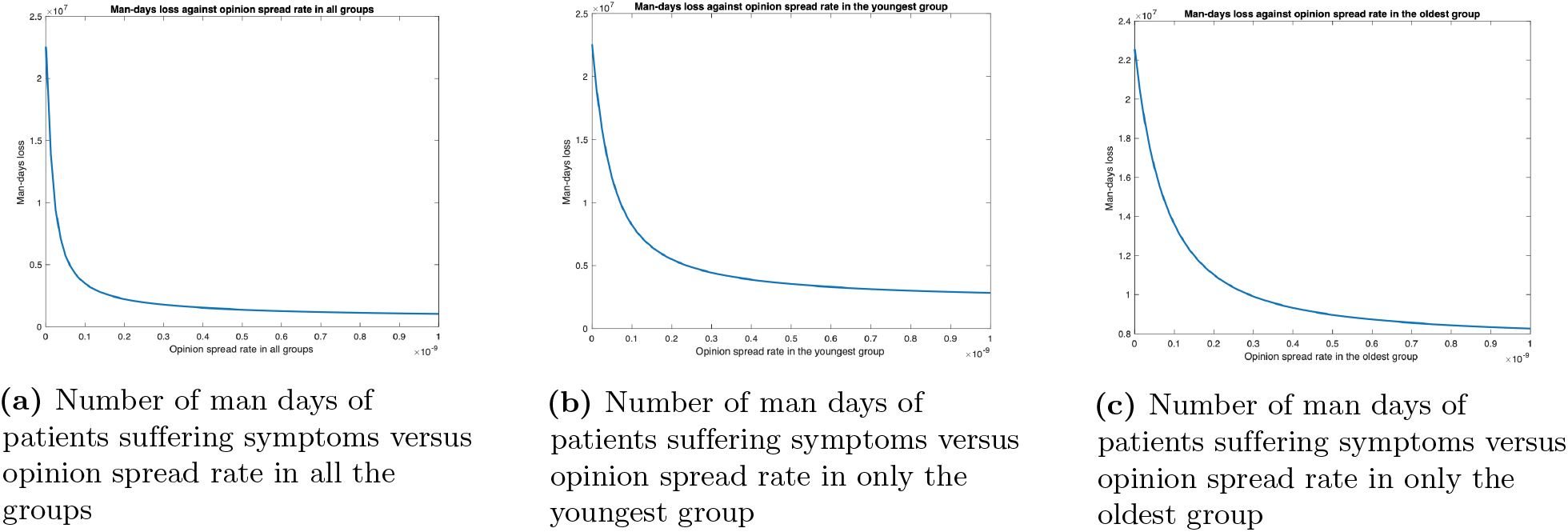
Impact of opinion dynamics on long COVID in Scenario 1

We observe that man days lost decreases as the opinion spread rate increases. The decrease is most when the opinion spread rate for all the groups increases, intermediate when the rate of spread of opinion increases only for the youngest group, and least when the rate of spread of opinion only increased for the oldest group. When the rate of spread of opinion for all the groups, only the youngest group, and only the oldest group increases, man-days lost due to long COVID decreases by 95.43%, 87.49%, and 63.39% respectively. Thus the impact of opinion dynamics is pronounced in this case as a higher rate of spread of opinion tantamounts to non-cooperatives converting to cooperatives at a higher rate, which in turn leads to a lower rate of spread of the disease in the populace and therefore lower number of man days individuals experience symptoms. Again the opinion dynamics in the youngest group have a much greater impact than that in the oldest group.

#### 3.2.4 Synthetic variant with high fatality rate

Next, we consider higher values of fatality rate motivated by the recent creation by BU researchers of a synthetic variant of COVID-19, namely the recombinant virus (Omi-S), that causes 80% fatality rate in mice [27]. Fig 11 show that fatality count increases by a factor of 8 as compared to when the fatality rate is at the default value (compare the numbers reported here with those in Fig 7). In addition, the decrease in fatality with the increase in the opinion spread rate follows the same pattern as in the default case. For instance, when the opinion spread rates for the youngest group increase from 0 to 2 *×* 10*^−^*^9^, the fatality count for all, youngest, middle age, and oldest groups respectively decrease by 88.22%, 90.27%, 88.36%, and 88.11% as against 88.22%, 90.43%, 88.35%, and 88.10% for the default case. Similarly, when the opinion spread rates for the oldest group increase from 0 to 2 *×* 10*^−^*^9^, the fatality count for all, youngest, middle age, and oldest groups respectively decreases by 69.24%, 64.82%, 64.89%, and 69.81% as against 69.23%, 64.89%, 64.77%, and 69.80% for the default case.

**Fig 11.**
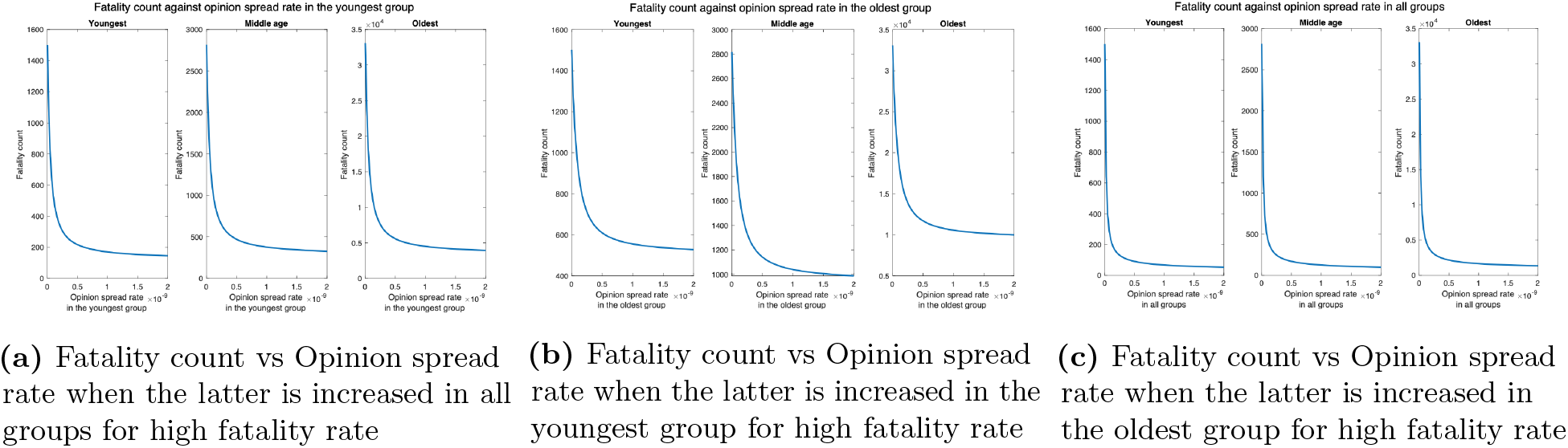
Synthetic variant with high fatality rate.

#### 3.2.5 When non-cooperatives convert cooperatives

We finally consider the case that non-cooperatives convert cooperatives while exchanging opinions. Now, as expected, unlike in the former case where cooperatives convert non-cooperatives, fatality increases with an increase in the opinion spread rate. We consider two values of the physical interaction rate in the youngest group: default value *y_L_* and thrice the default value *y_H_* (Fig 12). We observe that for *y_L_,* the fatality counts in all, youngest, middle age, and oldest groups respectively increase by 793.81%, 1136.36%, 866.67%, and 777.37% as the opinion spread rate in the youngest group increases from 0 through 2 *×* 10*^−^*^9^ (Fig 12a). For *y_H_* , the fatality count for all, youngest, middle age, and oldest groups will respectively increase by 48.21%, 51.11%, 43.28% and 48.37% as the opinion spread rate in the youngest group increases from 0 through 2 *×* 10*^−^*^9^. Similarly, for *y_L_,* the fatality count for all, youngest, middle age, and oldest group increase by 116.29%, 100.00%, 100.00% and 118.61% respectively, as the opinion spread rate of the oldest group increases from 0 through 2 *×* 10*^−^*^9^ (Fig 12b). For *y_H_* , the fatality counts for all, youngest, middle age, and oldest groups respectively increase by 30.61%, 5.83%, 12.22%, and 34.19% as the opinion exchange rate in the oldest group increases from 0 to 2 *×* 10*^−^*^9^. In addition, for *y_L_,* the fatality count for all, youngest, middle age, and oldest group increase by 5252.44%, 5600%, 5245.45%, and 5239.05% respectively, as the opinion spread rate increases from 0 through 2 *×* 10*^−^*^9^ in all groups (Fig 12c). Similarly, for *y_H_* the respective increase in this case are 115.93%, 59.39%, 107.70%, and 121.39%. We observe that as before, the highest increase in fatalities occurs when the opinion spread rate increase for all the groups, and if the opinion spread rate were to increase in only one age group, the increase in the youngest one has the maximum impact over all the groups.

**Fig 12.**
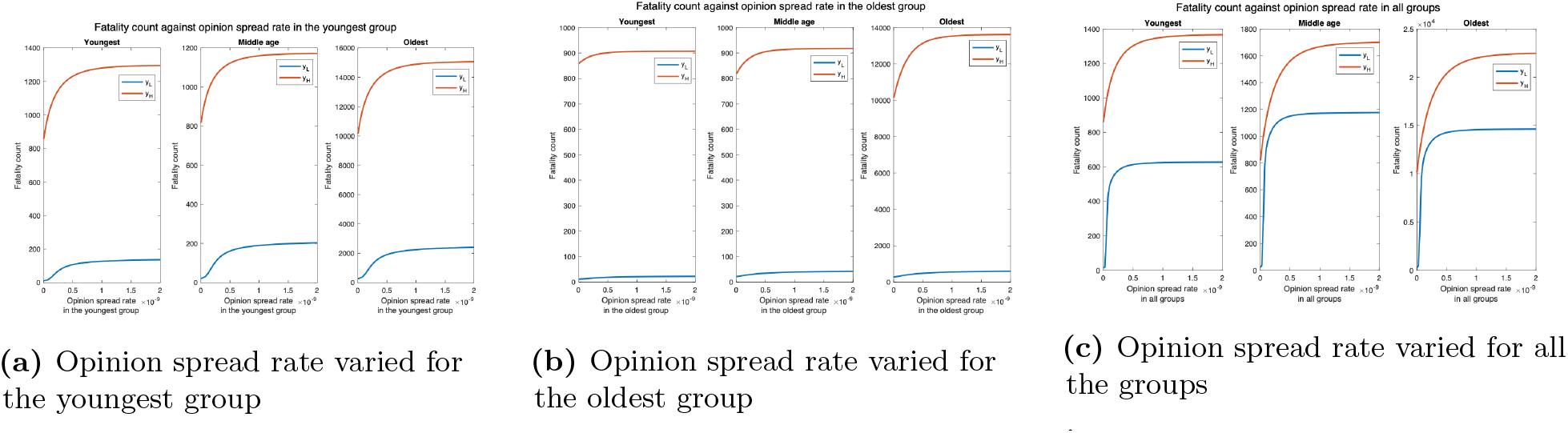
Non-cooperatives convert cooperatives during interactions. *y_L_* denotes the default interaction rates of the youngest group while *y_H_* represents when the youngest group interaction rates are tripled (other groups’ interaction rates remain the same).

### 3.3 Impact of opinion dynamics when behavioral practices affect the severity of symptoms of COVID-19: Scenario 2

We consider that cooperatives are willing to receive vaccines, but observe no other behavioral precaution; non-cooperatives refuse all behavioral precautions including receiving vaccines. We assumed that COVID-19 vaccination in the US is now readily available across the country and the vaccines need only one dose. Thus an individual is vaccinated right after he becomes willing to be vaccinated, i.e., cooperative. According to [44], the secondary attack rate (SAR) among household contacts exposed to fully vaccinated index cases was similar to household contacts exposed to unvaccinated index cases. Thus, vaccination does not affect the rate of transmission of the disease. Thus, cooperatives spread the disease at the same rate as non-cooperatives. However, it has been established that vaccines reduce the severity of symptoms, including hospitalization and death. For example, vaccines reduce non-intensive care unit (ICU) hospitalizations and deaths by 63.5% and 69.3% respectively [38]. Thus, infected cooperatives experience lower hospitalization and death rates as compared to infected non-cooperatives.

Since cooperatives transmit the disease at the same rate as non-cooperatives, conversion to cooperative does not affect the rate of spread of the disease. Thus, increasing the rate of spread of opinions in one age group affects the fatality count only in that group. Thus, as the rate of spread of opinion in the youngest group increases from 0 through 2 *×* 10*^−^*^9^, the fatality count for the middle age and oldest groups remain unchanged; the fatality count in the youngest group however decreases by 33.49% (Fig 13a). For *y_H_* , the disease spreads faster among the young and subsequently to the other groups. Thus, the fatality counts increase in all age groups in this case, but the fatality counts decrease with an increase in the opinion exchange rate only in the youngest group, the fatality counts do not change in other age groups. Similarly, when the opinion exchange rate increases only in the oldest group, the fatality count remains constant in other age groups but decreases by 33.47% in the oldest group (Fig 13b). Thus opinion dynamics in one group do not affect public health attributes among others.

**Fig 13.**
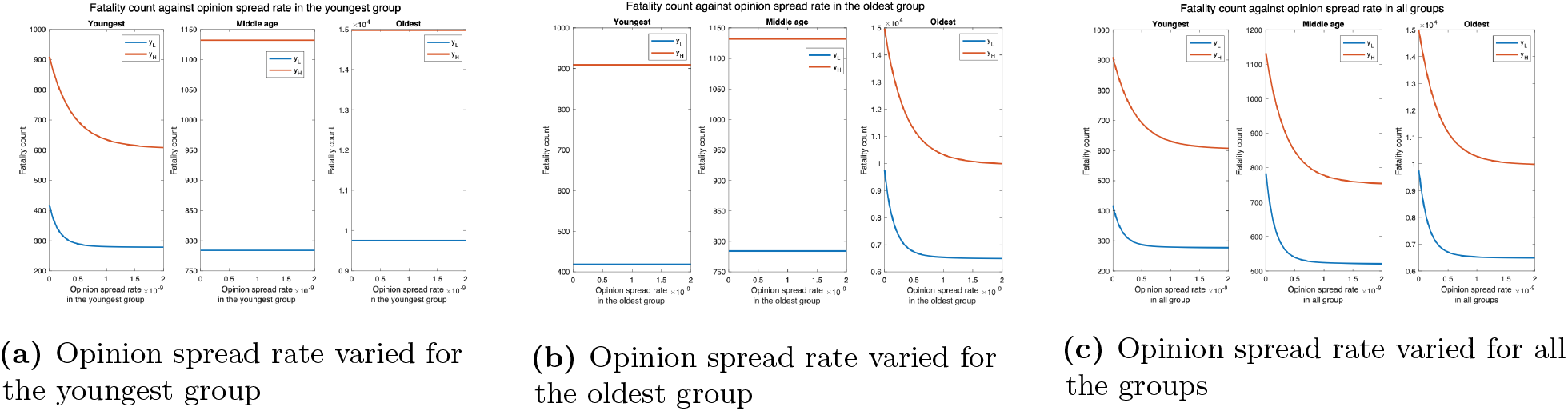
Impact of vaccination on fatality. *y_L_* denotes the default interaction rates of the youngest group while *y_H_* represents when the youngest group interaction rates are tripled (other groups’ interaction rates remain the same).

Next, we consider the impact of long COVID. For multidose vaccine, the prevalence of long COVID in vaccinated individuals is 30.0% after one dose, 17.4% after two doses, and 16.0% after a single dose. Since we consider only single-dose vaccines, we consider the overall prevalence to be the average, i.e. 21.13% [23, 24]. The number of man-days of symptoms can be computed similarly to that for Scenario 1, with the difference that *r*_1 ≠_ *r*_2_ in this case as different fractions of vaccinated and unvaccinated experience long COVID. The fraction of cooperatives and non-cooperatives who suffer long COVID are *r*_1_ = 0.2113 and *r*_2_ = 0.418 respectively [23, 24]. The man-days lost due to COVID decrease as the opinion spread rate increases (Fig 19). The decrease is most when the opinion spread rate increases in all groups, intermediate when the rate of spread of opinion increases only for the youngest group, and least when it increases only in the oldest group. When the rate of spread of opinion increases in all groups, only in the youngest group, and only in the oldest group, the number of man days decreases by 29.85%, 10.12%, and 9.09% respectively. In presence of long COVID, the man-days lost when cooperatives only accept vaccines is observed to be lower when compared to the case in which cooperatives observe the behavioral practices that impede COVID but don’t accept vaccines.

### 3.4 Impact of opinion dynamics when behavioral practices affect both spread and severity of symptoms of COVID-19: Scenario 3

We now consider that cooperatives follow behavioral practices that impede the spread of COVID-19 and reduce the severity of symptoms if one imbibes the disease (i.e., accepts vaccines). The fatality counts for all, youngest, middle age, and oldest groups respectively decrease by 88.24%, 93.18%, 88.31%, and 88.01% as the opinion spread rate of the youngest group increases from 0 through 2 *×* 10*^−^*^9^ (Fig 14a ). The fatality counts for all, youngest, middle age, and oldest group decrease by 78.55%, 64.39%, 64.52%, and 80.38% respectively, as the opinion spread rate of the oldest group improves from 0 through 2 *×* 10*^−^*^9^ (Fig 14b). Thus opinion exchange in all groups has the greatest impact, if opinions are to be exchanged in only one group, selecting the youngest group has the maximum impact. For *y_H_ ,* the number of deaths for all, youngest, middle age, and oldest groups will respectively decrease by 47.32%, 69.54%, 43.14%, and 49.36%, as the opinion spread rate in the youngest group is varied (Fig 14a). Similarly, the number of deaths for all, youngest, middle age, and oldest groups will respectively decrease by 43.60%, 2.66%, 6.64%, and 49.36%, as the opinion spread rate in the oldest group is varied (Fig 14b).

**Fig 14.**
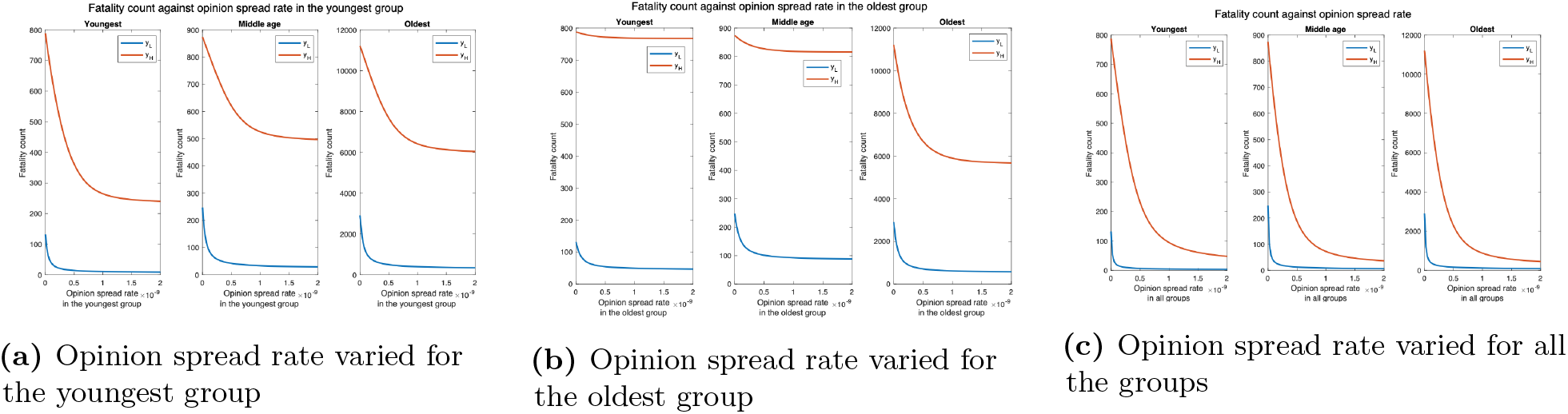
Joint implementation of vaccines and behavioral practices. *y_L_* denotes the default interaction rates of the youngest group while *y_H_* represents when the youngest group interaction rates are tripled (other groups’ interaction rates remain the same).

We now consider the impact of opinion dynamics in presence of long COVID. The number of man-days lost due to COVID-19 decrease as the opinion spread rate increases (Fig 20). The decrease is most when the opinion spread rate for all the groups improve, intermediate when the rate of spread of opinion increases only for the youngest group, and least when the rate of spread of opinion only increases for the oldest group. When the rate of spread of opinion for all the groups, the youngest group only, and oldest group only increases from 0 through 2 *×* 10*^−^*^9^, the above attribute decreases by 95.98%, 88.18%, and 65.50% respectively. These decreases are considerably higher compared to that when cooperatives only accept vaccines and do not adopt other behavioral precautions; the decreases are only 29.85%, 10.12%, and 9.09% in this case (Scenario 2, Section 3.3). This is because we had considered that vaccines do not reduce the rate of transmission of disease (based on cited literature). If vaccines reduced transmission of the disease as well, at the same rate as behavioral practices, the results in this section will be obtained even when cooperatives only accept vaccines.

## 4 Discussion

We summarize the important findings from our work and the implications thereof to practice. We have presented a mathematical model that can be easily adapted to model the impact of a wide range of behaviors on public health attributes during the spread of COVID-19. Our model is therefore flexible, and also computationally tractable in that the size of the population does not affect the computation time. Thus the model can scale to large population sizes which is an important strength given that the target population for a pandemic is typically large and easily in the order of millions considering populations of large cities and even mid-size provinces. Some of our findings from the model reinforce prior expectations by quantifying the impact. For example, we show that following a combination of behavioral practices that impede the spread of COVID-19 and partaking in vaccines that reduce the severity of symptoms together substantially reduce both the fatalities and the number of man-days lost due to symptoms from long COVID. This conclusion is only expected but our findings quantify the improvements that accompany good health practices. Next, we show that opinion dynamics pertaining to the observance of behavioral practices that impede the spread have a substantial impact on fatality count and man-days lost due to symptoms. In particular, a higher rate of exchange of opinions that convert (respectively dissuade) individuals to observing such practices substantially decreases (respectively increases) the fatality count and man-days lost due to symptoms. Our work is the first to quantify the impact of behavioral dynamics and the spread of opinions pertaining to the observance of desired behaviors on public health damages inflicted by COVID-19. We also show that opinion dynamics in groups that have the highest rate of physical interaction have the maximum impact on overall fatalities and man-days lost due to symptoms even though such groups are least likely to imbibe severe forms of the disease. Specifically, if the rate of spread of opinion that conforms to desirable behavioral practices increases among the youngest groups, fatality for all the groups decreases. If the rate of spread of similar opinions increases among the oldest age group, then the positive impact due to the spread is largely confined to the same age group. We show that this disparity arises because young individuals physically interact among themselves and across age groups at a higher rate than others. The consequence of this finding is that it reveals how mal-actors can undermine public health in society by deliberately targeting young individuals with a propaganda campaign that dissuades them from observing desirable health practices. Note that such dissuading may be relatively simple because their own risk of imbibing a severe form of the disease is low. Appropriate precautions involving a counter-information campaign can be undertaken once this vulnerability is identified and its impact quantified which our work has accomplished.

In conclusion, we discuss a generalization of our model. We have so far assumed that vaccines require only one dose. But, according to CDC, there are four approved or authorized vaccines in the United States namely Pfizer-BioNTech, Moderna, Johnson & Johnson’s Janssen, and Novavax COVID-19 vaccines; and Johnson & Johnson’s Janssen requires one dose but all the rest require two doses three to four weeks apart. We now describe how our model can be generalized to accommodate two doses. Refer to Fig 15 for a pictorial illustration of the state diagram that captures this generalization while considering classification based only on opinions. The only difference with the previous versions (i.e., Fig 3) is that we introduce a new vaccinated state (*V* ) and cooperative susceptibles and exposed transition to this state after they receive the second dose. When an individual becomes cooperative we consider that he has received the first dose because currently in many countries the first dose can be received without any administering delay. The transition time between the cooperative and vaccinated state is the delay mandated between the two doses. According to the CDC, people who tested positive for COVID-19 (symptomatic or asymptomatic) should wait to be vaccinated until they have recovered from their illness and have met the criteria for discontinuing isolation [3]. We, therefore, do not include any transition from symptomatic or asymptomatic states to vaccinated. Thus a vaccinated individual can get infected and transition to the exposed state *E_v_,* then to either the presymptomatic or asymptomatic stage at a certain probability. The asymptomatic vaccinated recovers after some days while the presymptomatic vaccinated experiences symptoms after about 2 days. Thereafter, the symptomatic vaccinated either get hospitalized at a reduced hospitalization rate or recover. The hospitalized individual eventually recovers or dies. The hospitalization and fatality rates for vaccinated individuals are much lower compared to those for unvaccinated individuals. These can be incorporated by appropriately choosing the transition probabilities from the vaccinated state.

**Fig 15.**
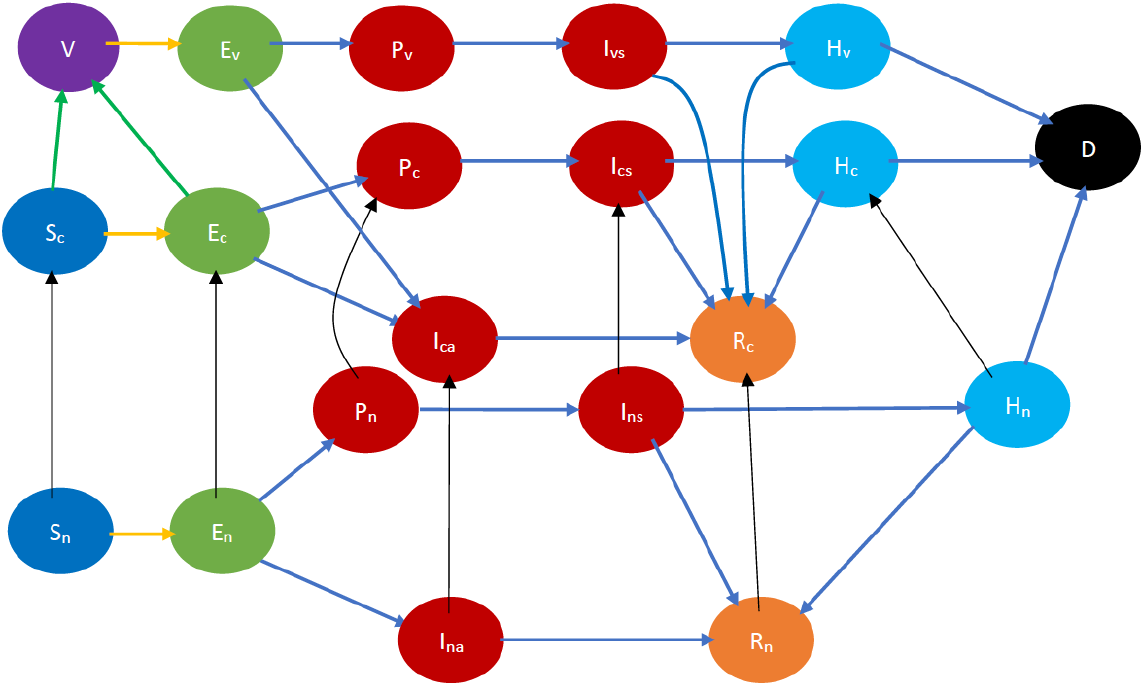
COVID-19 state diagram for two-dose vaccine. The state in purple color is the vaccinated state. The states in blue color are the susceptibles while those in light green color are exposed (still in the incubation period). The states in dark red are infected and therefore infectious while those in gold have recovered, those in light blue are hospitalized, and black denotes dead. In addition, the green arrows denote transition to the vaccinated state after receiving the vaccine, the yellow arrows show susceptibles transitioning to the exposed state after contracting the virus, the black arrows indicate opinion evolution, and the blue arrows indicate the natural progression of the disease. The yellow arrows show susceptibles transitioning to the exposed state after contracting the virus.

## Data Availability

All data produced in the present work are either contained in the manuscript or available online (reference in the present work) at 23andMe.

https://you.23andme.com/covid19/

## Supporting information I

### Equations of SARS-CoV-2 disease dynamics

#### Developing the Clustered Epidemiological Differential Equations (CEDE) for COVID-19

Table 1 contains all the fundamental states in our model while Table 2 contains all SARS-CoV-2 transition parameters. Referring to the notations in Table 1, we index each term by the group the individual in the state belongs. More specifically, *S_c_*(*t*) is the fraction of individuals who are susceptible and cooperative at time *t. S_n_*(*t*)*, E_c_*(*t*)*, E_n_*(*t*)*, P_c_*(*t*)*, P_n_*(*t*)*, I_cs_*(*t*)*, I_ns_*(*t*)*, I_ca_*(*t*)*, I_na_*(*t*)*, H_c_*(*t*)*, H_n_*(*t*) may be defined similarly using Table 1 as a basis. Finally, *R_c_*(*t*)*, R_n_*(*t*) are respectively the fractions of cooperative and non-cooperative individuals who recover from COVID-19 at time *t* while *D*(*t*) denotes those that died due to COVID-19 at time *t.* Let *I*(*t*) be the fraction of the *infectious* individuals at time *t.* That is, *I*(*t*) is the fraction of individuals who can infect susceptible individuals through contact. Then,

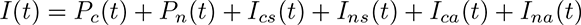

**Table 2.** SARS-CoV-2 transition parameters

| Parameters | Description of parameters |
| --- | --- |
| $\frac{1}{\lambda}$ | Expected time an exposed individual is in presymptomatic stage |
| $\frac{1}{\gamma}$ | Expected time an individual is in presymptomatic stage |
| $\frac{1}{\mu}$ | Expected time a symptomatic individual is in infection stage |
| $\frac{1}{\beta}$ | Expected time an asymptomatic individual is infectious |
| $\frac{1}{\mu}$ | Expected time a symptomatic individual is in late infection stage |
| $\frac{1}{\sigma}$ | Expected time an individual hospitalized |
| $p$ | Probability that an infected individual will be asymptomatic |
| $\eta$ | Probability that an individual at late infection stage is hospitalized |
| $\lambda$ | Probability that an individual at late infection stage dies |

Interaction between individuals corresponding to *S_c_*(*t*)*, S_n_*(*t*) and *I*(*t*) spread the disease to the susceptibles and transform them to Exposed states *E_c_*(*t*)*, E_n_*(*t*) - refer to the yellow arrows in Fig 4. The natural progression of the disease changes *E_c_*(*t*) to either *P_c_*(*t*) or *I_ca_*(*t*) at a certain probability. Similarly, with a certain probability, *E_n_*(*t*) transitions to either *P_n_*(*t*) or *I_na_*(*t*)*. P_c_*(*t*) would then transition to *I_cs_*(*t*), and then to either *H_c_*(*t*) or *R_c_*(*t*) or *D*(*t*) while *P_n_*(*t*) would then transition to *I_ns_*(*t*) to either *H_n_*(*t*) or *R_n_*(*t*) or *D*(*t*). In addition, *I_ca_*(*t*) and *I_na_*(*t*) respectively transition to *R_c_*(*t*) and *R_n_*(*t*). Finally, *H_c_*(*t*) and *H_n_*(*t*) transition to either *R_c_*(*t*) or *R_n_*(*t*) or *D*(*t*) - refer to the blue arrows in Fig 4. The transitions are similar for the males and females, different age groups and races, healthy individuals, and individuals with underlying health conditions, but the fractions of those who recover in each populace are different.

Individuals in any group can get in physical proximity to one another at a certain rate. The rate is much higher if the individuals are of the same race and within the same age group. Meanwhile, only a fraction of these contacts spread the disease. We consider that the disease spread rate is the product of the rate at which individuals are in physical contact with one another and the fraction of such contacts between susceptibles and infectious individuals. The physical contact rates of the system are also known as the disease spread rates. The contact rate differs for the various age groups. We use *ϕ* to denote the physical rate of interaction between individuals of the same race and age group (youngest and middle age groups). Older people have smaller, more family-centric networks, and spend less time with others [37]. Thus, their rate of interaction is lower compared to the other groups. Therefore, we denote the disease spread rate between the oldest group of the same race with *mϕ* where *m <* 1. Similarly, we denote the physical contact rate between the oldest group and the other groups with *κϕ* where *κ <* 1. In addition, we represent the physical contact rate between the youngest age group of different races with *aϕ,* middle age group of different races as *bϕ,* and oldest age group of different races with *cϕ.* We denote any other physical interaction rate with *xϕ,* where *a <* 1*, b <* 1*, c <* 1*, x <* 1. Similarly, we use *α* to represent the opinion spread rate. In this case, ideas are exchanged during interactions - which might lead to a change in opinion. Meanwhile, one may be converted by exposure to public awareness campaigns (which may have a stronger impact because the individual might be infected and experiences the symptoms acutely). We denote the rate of such opinion change as *δ*.

Interaction between susceptible individuals i.e. *S_c_*(*t*)*, S_n_*(*t*) and Infectious individuals i.e. *P_c_*(*t*)*, P_n_*(*t*)*, I_cs_*(*t*)*, I_ns_*(*t*)*, I_ca_*(*t*)*, I_na_*(*t*) may spread the disease to the susceptibles and transform them to early incubators - refer to the yellow arrows in Fig 4. We assumed that those that are hospitalized, *H_c_*(*t*)*, H_n_*(*t*), are isolated from the general public (quarantined) and hence cannot infect other susceptible nor can they convert the opinions of others. Natural progression of the disease change *E_c_*(*t*) to *P_c_*(*t*) to *I_cs_*(*t*) to either *H_c_*(*t*) or *R_c_*(*t*) or *D*(*t*); also, *E_c_*(*t*) to *I_ca_*(*t*) to *R_c_*(*t*) - refer to the blue arrows in Fig 4. The transitions due to the natural progression of the disease are similar for cooperatives and non-cooperatives.

We model the evolution of the states through a set of meta-population epidemiological differential equations. Each differential equation captures the evolution of a particular variable. Thus, the solution of the system of differential equations provides the fraction of individuals in different states at given times, that is, the spatio-temporal distribution of the disease and opinion spread. The terms in the differential equations are either quadratic or linear. The quadratic ones represent the transitions brought on by interactions between two individuals, specifically physical proximity, and exchange of ideas (refer to the yellow arrows and the black arrows in Fig 4) and the linear ones represent the transitions that happen otherwise, specifically natural progression of the disease, conversion through reading, media, etc. (refer to the blue and black arrows in Fig 4). Note that interactions always involve two individuals, hence interactional transitions are represented by quadratic terms; in contrast, the non-interactional transitions involve only one individual and are therefore represented by linear terms.

Let *I*(*t*) be the fraction of the infectious individuals at time *t* and *X*(*t*) be the fraction of individuals who are cooperative at time *t.* The resulting clustered epidemiological differential equations (CEDE) are given in equations (1) - (15).

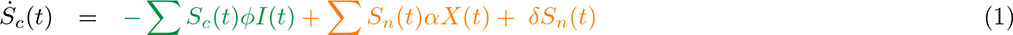

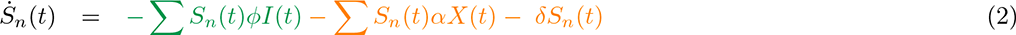

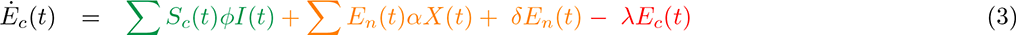

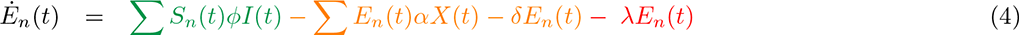

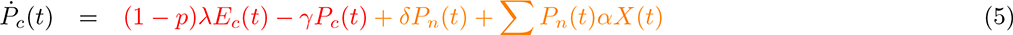

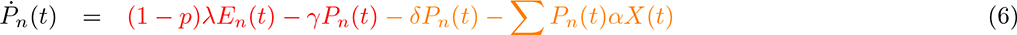

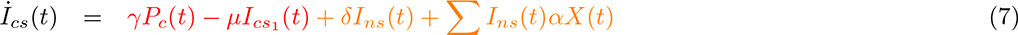

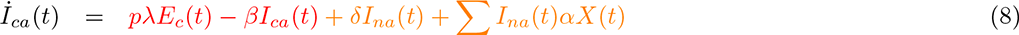

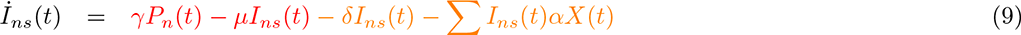

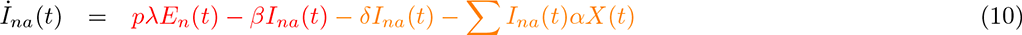

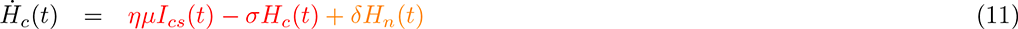

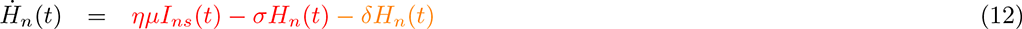

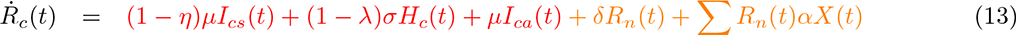

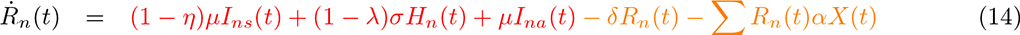

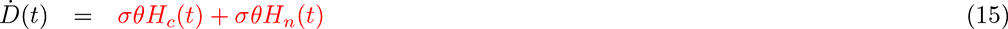

The terms in green color are quadratic terms that represent the spread of the SARS-CoV-2 to the susceptibles due to physical interaction with the infectious individuals and the subsequent transformation of the susceptibles to the exposed states - refer to the yellow arrows in Fig 4. The terms in orange represent the transformation of non-cooperative individuals to cooperative ones - refer to the black arrows in Fig 4. The quadratic terms represent those conversions due to the exchange of opinions during interaction whereas the linear terms represent changes in opinion due to public awareness campaigns. The terms in red represent the natural progression of the disease - refer to the blue arrows in Fig 4. The differential equations for the evolution of COVID-19 in cooperatives are identical to those of the non-cooperatives, as can be seen in equations 1 to 15.

Note that (1) is similar to (2) as transitions are similar for the cooperative and non-cooperative individuals. The same observation may be made for ((3), (4)), ((5), (6)), ((7), (8)), ((9), (10)), ((11), (12)) , ((13), (14)) . So, we only explain (1), (3), (5), (7), (9), (11), (13), and (15).

The first terms in (1), (3), (the terms in green color) are quadratic terms that represent the spread of the disease to the susceptibles due to interaction with the infectious individuals and the subsequent transformation of the susceptibles to early incubators - refer to the yellow arrows in Fig 4. The rate of conversion of susceptibles in a group to the exposed stage is proportional to the number of physical contacts per unit time between the susceptibles with infectious individuals since each such contact spreads the disease to the susceptible with a certain probability. The proportionality constant here is the probability that such a contact spreads the disease or equivalently the fraction of such contacts that spread the disease. The number of such physical contacts per unit time again is proportional to the number of pairs of susceptibles in the group in question and infectious in the same and other groups. Here the proportionality constant is the reciprocal of the expected time between successive physical contacts between individuals in a given such pair. The expected time will presumably be lower, and therefore the proportionality constant higher, if both individuals are in the same group than if they are in different groups since connections between individuals of the same age, race, and gender are usually more frequent [30, 37, 43]. The number of pairs of individuals one of which is susceptible in one group and another an infectious individual in the same or other groups is the product of the number of susceptibles in one group and infectious in the same or another group. These products lead to the quadratic terms. The overall proportionality constants are the products of the two proportionality constants mentioned above, and give us *ϕ*, the disease spread rates, which have been summarized in Table 6. Since the spread of the disease reduces the number of susceptibles and increases the number of early incubators, the first term in (1) has a positive sign and that in (3) has a negative sign.

The second terms in (1) through (14) (the terms in orange) are quadratic terms that represent the transformation of non-cooperative individuals to cooperative ones through the exchange of opinions with cooperative individuals - refer to the black arrows in Fig 4. The rate of transformation for non-cooperatives of a certain type (the type is specified by the stage of disease, cooperativity, health status, gender, race, and age) is proportional to the number of opinion exchanges per unit time between the non-cooperatives and cooperatives; the proportionality constant is the value of the above fraction. The number of such exchanges per unit time again is proportional to the number of pairs of non-cooperatives of the type in question and cooperatives in the same and other groups. The number of pairs of individuals one of which is a non-cooperative of a certain type and another a cooperative in the same or another group is the product of the number of non-cooperatives of the type and cooperatives in the same or other groups. These products lead to the quadratic terms. The overall proportionality constant is the product of the two proportionality constants above, and provides us the opinion spread rates, *α.* The third terms in (1) through (14) (the terms in orange) are linear terms that represent the transformation of non-cooperative individuals to cooperative ones through exposure to public awareness campaigns (which may have a stronger impact because the individual might be infected and thus experiences the symptoms acutely). We denote the rate of such opinion change as *δ*. Since the change of opinion increases (respectively decreases) the cooperatives (respectively non-cooperatives), the second terms in the equations for the cooperatives (e.g., (1)) are positive and the second terms for the non-cooperatives (e.g., (2)) are negative.

All other terms in (3), (5), (7), (9), (11), (13), and (14) (the terms in red) represent the natural progression of the disease - refer to the yellow arrows in Fig 4. For instance, the second term in (5) and the first term in (7) represent the natural progression from the presymptomatic to the symptomatic infectious state. The first term in (13) represents the recovery of cooperative individuals from the symptomatic infected stage. Finally, the solution of the CEDE provides the spatio-temporal distribution for the spread of the disease, namely the fraction of individuals who (1) are dead in any group at time *t* (*D*(*t*)); (2) have recovered in any group at time *t* (*R*(*t*)); (3) are susceptible and are in any group at time *t* ((*Sc* + *S_h_*)(*t*)); (4) are infected in any group at time *t* ((*P_ci_* + *P_hi_* + *C_ci_* + *C_hi_* + *E_ci_* + *E_hi_*)(*t*)); (5) are contagious and are in cluster *i* at time *t* ((*P_c_* + *P_n_* + *I_cs_* + *I_ns_* + *I_ca_* + *I_na_*)(*t*)).

Meanwhile, equations (1) - (15) represent the basic compartments in our model. We now generalize the states in our model. Recall that an individual is characterized by his cooperativity, health conditions, gender, ancestry, age, stage of the disease, as well as symptomatic or asymptomatic manifestation. Thus, we use the suffixes *c* to denote cooperativity and *n* to denote non-cooperativity. Similarly, *h* denotes being healthy (immunocompetent), *d* denotes immunodeficient (not healthy, immunocompromised), *m* represents male, while *f* represents female. In addition, we use the subscripts *x*, *y*, and *z* respectively to represent individuals of African American, Hispanic, and White-American ancestry. Furthermore, we classify the age of an individual into three groups with subscript 1 denoting 0 *−* 24 years, 2 denoting 25 *−* 49 years, and 3 denoting 50 years and above. Finally, we use the suffixes *s* and *a* to respectively denote symptomatic and asymptomatic individuals. Therefore, the state *S_chmx_*_1_ denotes cooperative, healthy, male susceptible African American in the 0 *−* 24 years age group. Similarly, *E_ndfx_*_3_ denotes non-cooperative, not healthy (immunocompromised), exposed female Hispanic/Latino who is 50 years and above, *I_chmzs_*_2_ denotes cooperative, healthy, infected symptomatic male White-American in the 25 *−* 49 years age group while *I_ndfza_*_1_ denotes non-cooperative, not healthy (immunocompromised), infected asymptomatic female African American in the 0 *−* 24 years age group.

## Supporting information II

### Parameter estimation

Table 3 shows the demographic characteristics of Pennsylvania with respect to age and race [9]. Note that few percentages of the population are neither African Americans, Hispanics/Latinos, nor White-American. Thus, we distributed such population according to the percentage of those 3 races we considered. For instance, for the age group 25 - 49 years, there are 4.4% African Americans, 1.3% Hispanics, 24.0% White-American, and 2.0% other races. We distributed the 2.0% population thus: African Americans = 4.4 + (4.4 *×* 2)/29.7 = 4.7%. Similarly, Hispanics = 1.3 + (1.3 *×* 2)/29.7 = 1.4%, while White-American = 24.0 + (24 *×* 2)/29.7 = 25.6%. In addition, the gender distribution in Pennsylvania is 51.1% female and 48.9% male [13]. The total number of people we considered in our model is 10 million whereas our choice for the initial number of infected individuals is 10000.

**Table 3.**
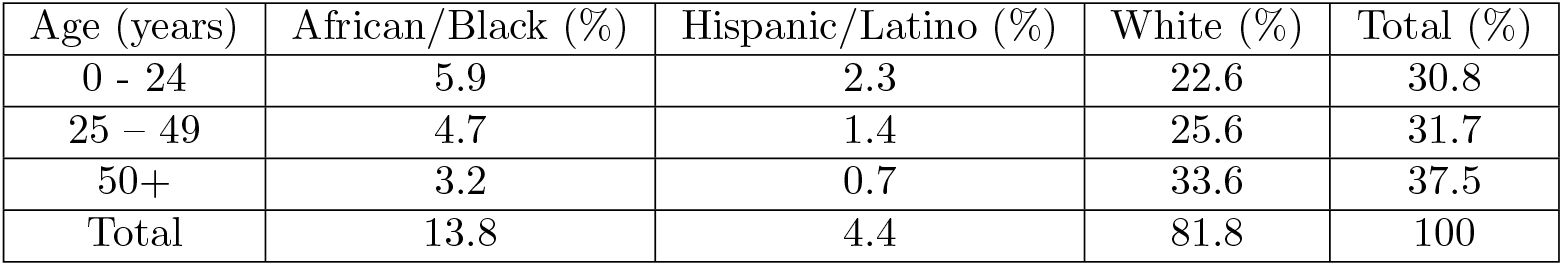
Population distribution by race and age.

Comorbidity refers to the existence of more than one disease or condition in the same person at the same time. The Centers for Disease Control and Prevention (CDC) and the PA Department of Health (PaDOH) have highlighted concern for individuals of all ages who suffer from underlining medical conditions such as obesity, as well as those who are immunocompromised due to conditions like cancer treatment and being HIV/AIDS positive [26]. Furthermore, according to [39], about 40% of the US population are immunocompromised. Body Mass Index (BMI) is a person’s weight in kilograms divided by the square of height in meters. BMI over 30 is considered obese [4]. The average obesity rate among African American/Black, Hispanic/Latino, and White in Pennsylvania are 41.8%, 32.9%, and 31.3% respectively [21]. In this paper, we refer to people without any comorbidities as healthy. We obtain the fraction of people with underlying medical conditions by taking the average of immunocompromised and obese individuals. Thus, comorbidity for Africa America is (0.2 + 0.418)/2 = 0.3090; Hispanic = (0.2 + 0.329)/2 = 0.2645; and White = (0.2 + 0.313)/2 = 0.2565.

According to the Centers for Disease Control and Prevention (CDC), the current best estimate of the basic reproduction number, *R*_0_ for COVID-19 is 2.5 [8]. Recall that *R*_0_ is the average number of secondary infections caused by a single typical infected individual among a completely susceptible population. If *R*_0_ *>* 1, epidemic takes off. On the other hand, if *R*_0_ *<* 1, no major epidemic occurs. In addition, following the procedure outlined in (Chapter 6, [41]), we obtain the expression for *R*_0_ for our model as shown in equation 16.

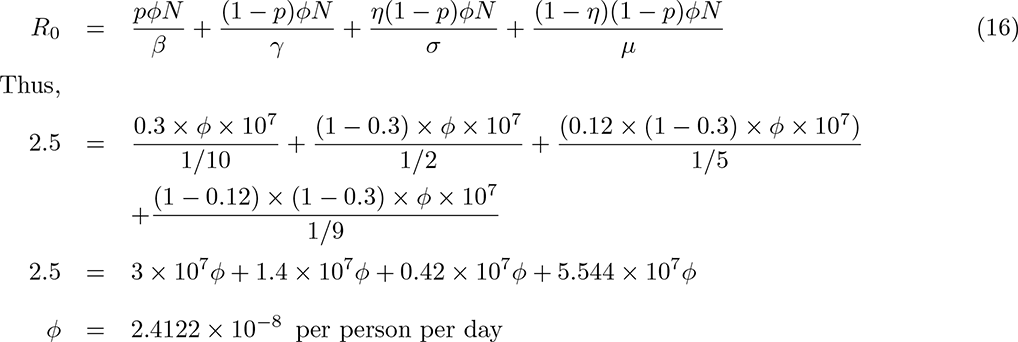

Furthermore, 30% of infected individuals are asymptomatic [8]. The transmission of SARS-CoV-2 from an infected person to a secondary patient before the source patient developed symptoms is known as presymptomatic transmission [15]. The presymptomatic stage lasts for 2 days on average [10], [15]. The mean time from exposure to symptom onset is 6 days [8], i.e., 1*/λ* + 1*/γ* = 6 days. But 1*/γ* = 2 days on average [10]. Therefore, 1*/λ* = 4 days, and 1*/γ* = 2 days. The median number of days from symptom onset to hospitalization is 5 days [8]. That is expected time 1*/ϕ* + 1*/β* = 5 days. Therefore, 1*/β* = 2 days. The median number of days from symptom onset to death is 15 days [8]. Therefore, 1*/σ* = 15 - 1*/µ −* 1*/β* = 15 – 3 – 3 = 9 days. Thus, 1*/σ* = 9 days. Furthermore, the median duration of hospital stays among survivors = 9.3 days [36]. Similarly, according to CDC, the persons who never develop symptoms, isolation, and other precautions can be discontinued 10 days after the date of their first positive RT-PCR test for SARS-CoV-2 RNA. Therefore, 1*/β* = 10 days [5, 11]. The percent that dies among those hospitalized is 0.7% (0 – 17 years old), 2.1% (18 – 49 years old), 7.9% (50 – 64 years old), 18.8% (*≥* 65 years old) [8]. Therefore, fatality rates among hospitalized patients are: (0.7 + 2.1)*/*2 = 1.4% for 0 – 24 years, 2.1% for 25 – 49 years, and (7.9 + 18.8)*/*2 = 13.35% for 51+ years. Similar to [20], we define the fractions of individuals who fully comply with COVID-19 prevention measures at the initial time as the *initial cooperativity*. According to [39], approximately 40% of the populace is not cooperative. Thus, our default choice for the initial cooperativity is 0.6, but we also consider other values of initial cooperativity. We assume that interaction between a pair of non-cooperatives is twice likely to result in infection. In addition, we assume that the disease spread rate between a pair of individuals such that only one is non-cooperative equals that when both are non-cooperatives. COVID-19 vaccines have been shown to be effective and help to reduce hospitalizations, intensive care unit admissions, and deaths [18, 42]. However, the secondary attack rate among household contacts exposed to fully vaccinated index cases was similar to household contacts exposed to unvaccinated index cases [40]. Meanwhile, vaccination against COVID-19 is now seamless and vaccines are readily available to receive across the U.S. Thus, we also considered cases in which non-cooperatives are assumed to be twice likely to be hospitalized compared with cooperatives. The parameters we use in this research alongside their estimated values are outlined in Table 4.

**Table 4.** Parameter estimation

| Parameter | Description | Value | Ref. |
| --- | --- | --- | --- |
| $p$ | Probability that an infected person is asymptomatic | 0.3 | [8] |
| $\eta$ | Probability that an infected person is hospitalized | Table 5 | [1] |
| $\lambda$ | COVID-19 fatality rate | see p. 38 | [8] |
| $\frac{1}{\lambda}$ | Expected time an individual is in exposed stage | 4 days | p. 38 |
| $\frac{1}{\gamma}$ | Expected time an individual is pre-symptomatic | 2 days | [10, 15] |
| $\frac{1}{\mu}$ | Expected time an individual is infected before hospitalization | 5 days | [8] |
| $\frac{1}{\sigma}$ | Expected time for a hospitalized individual to recovers | 9 days | [36], p. 38 |
| $\frac{1}{\mu}$ | Expected time for an infected asymptomatic individual to recover | 10 days | [5, 11] |
| $R_0$ | Basic Reproduction number | 2.5 | [8] |
| $\phi$ | Disease spread rate | $2.4122 \times 10^{-8}$ | p. 38 |

**Table 5.** The average probability of hospitalization [1].

| Age<br>(years) | Sex | Health<br>condition | Probability of hospitalization |  |  |
| --- | --- | --- | --- | --- | --- |
|  |  |  | Africa Americans | Hispanics | Whites |
| 0 - 24s | M | Healthy | 0.03 | 0.03 | 0.03 |
| 0 - 24s | F | Healthy | 0.03 | 0.03 | 0.03 |
| 0 - 24s | M | Comorbid | 0.19 | 0.14 | 0.12 |
| 0 - 24s | F | Comorbid | 0.16 | 0.11 | 0.09 |
| 25 - 49s | M | Healthy | 0.05 | 0.03 | 0.03 |
| 25 - 49s | F | Healthy | 0.04 | 0.03 | 0.03 |
| 25 - 49s | M | Comorbid | 0.25 | 0.18 | 0.16 |
| 25 - 49s | F | Comorbid | 0.21 | 0.15 | 0.13 |
| 50+ | M | Healthy | 0.15 | 0.10 | 0.09 |
| 50+ | F | Healthy | 0.12 | 0.08 | 0.08 |
| 50+ | M | Comorbid | 0.50 | 0.40 | 0.36 |
| 50+ | F | Comorbid | 0.44 | 0.35 | 0.31 |

**Table 6.** Interaction rates.

|  | X1 | X2 | X3 | Y1 | Y2 | Y3 | Z1 | Z2 | Z3 |
| --- | --- | --- | --- | --- | --- | --- | --- | --- | --- |
| X1 | $\phi$ | $\phi$ | $k\phi$ | $a\phi$ | $x\phi$ | $x\phi$ | $a\phi$ | $x\phi$ | $x\phi$ |
| X2 | $\phi$ | $\phi$ | $k\phi$ | $x\phi$ | $b\phi$ | $x\phi$ | $x\phi$ | $b\phi$ | $x\phi$ |
| X3 | $k\phi$ | $k\phi$ | $m\phi$ | $x\phi$ | $x\phi$ | $c\phi$ | $x\phi$ | $x\phi$ | $c\phi$ |
| Y1 | $a\phi$ | $x\phi$ | $x\phi$ | $\phi$ | $\phi$ | $k\phi$ | $a\phi$ | $x\phi$ | $x\phi$ |
| Y2 | $x\phi$ | $b\phi$ | $x\phi$ | $\phi$ | $\phi$ | $k\phi$ | $x\phi$ | $b\phi$ | $x\phi$ |
| Y3 | $x\phi$ | $x\phi$ | $c\phi$ | $k\phi$ | $k\phi$ | $m\phi$ | $x\phi$ | $x\phi$ | $c\phi$ |
| Z1 | $a\phi$ | $x\phi$ | $x\phi$ | $a\phi$ | $x\phi$ | $x\phi$ | $\phi$ | $\phi$ | $k\phi$ |
| Z2 | $x\phi$ | $b\phi$ | $x\phi$ | $x\phi$ | $b\phi$ | $x\phi$ | $\phi$ | $\phi$ | $k\phi$ |
| Z3 | $x\phi$ | $x\phi$ | $c\phi$ | $x\phi$ | $x\phi$ | $c\phi$ | $k\phi$ | $k\phi$ | $m\phi$ |

Next, we use data from [1] to estimate the probability of hospitalization for African Americans, Hispanics/Latinos, and White Americans for the various age groups. We assumed that each individual averages 1 – 4 minutes of exercise per day. We also assumed that a healthy individual weighs 147 lbs on average while an obese individual weighs 203 lbs or more.

We use the subscripts *x, y,* and *z* to respectively represent people of African American, Hispanic, and White American ancestry. We also classify the age of an individual into three groups with subscript 1 denoting 0 *−* 24 years, 2 denoting 25 *−* 49 years, and 3 denoting 50 years and above. We use *ϕ* to denote the physical rate of interaction between individuals of the same race and age group (youngest and middle age groups). Older people have smaller, more family-centric networks, and spend less time with others [37]. Thus, their rate of interaction is lower compared to the other groups. Therefore, we denote the disease spread rate between the oldest group of the same race with *mϕ* where *m <* 1. Similarly, we denote the physical contact rate between the oldest group and the other groups with *κϕ* where *κ <* 1. In addition, we represent the physical contact rate between the youngest age group of different races with *aϕ,* middle age group of different races as *bϕ,* and oldest age group of different races with *cϕ.* We denote any other physical interaction rate with *xϕ,* where *a <* 1*, b <* 1*, c <* 1, and *x <* 1. As shown above, *ϕ* = 2.4122 *×* 10*^−^*^8^ per person per day, *a* = 0.9*, b* = 0.8*, c* = 0.2*, k* = 0.6*, x* = 0.3, and *m* = 0.4. Similarly, we use *α* to represent the virtual interaction rate also known as the opinion spread rate. We assumed that *α* = 10*^−^*^9^ and *δ* = 2 *×* 10*^−^*^9^ (default values).

## Supporting Information III

### Supplementary figures

**Fig 16.**
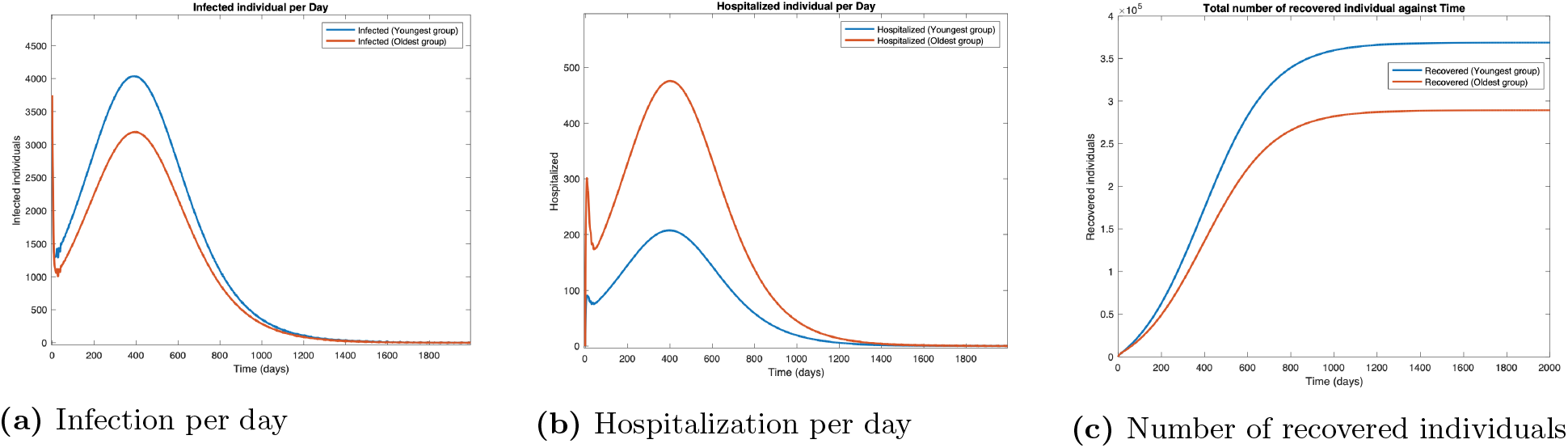
Scenario 1. Plots in the time domain for sanity check.

**Fig 17.**
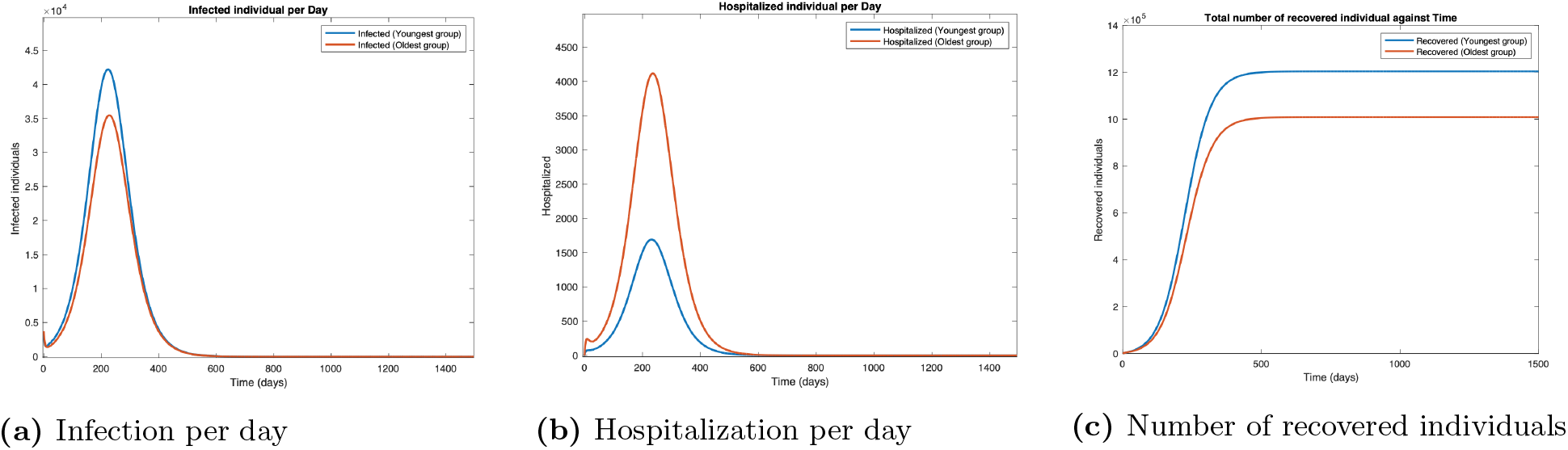
Scenario 2. Plots in the time domain for sanity check.

**Fig 18.**
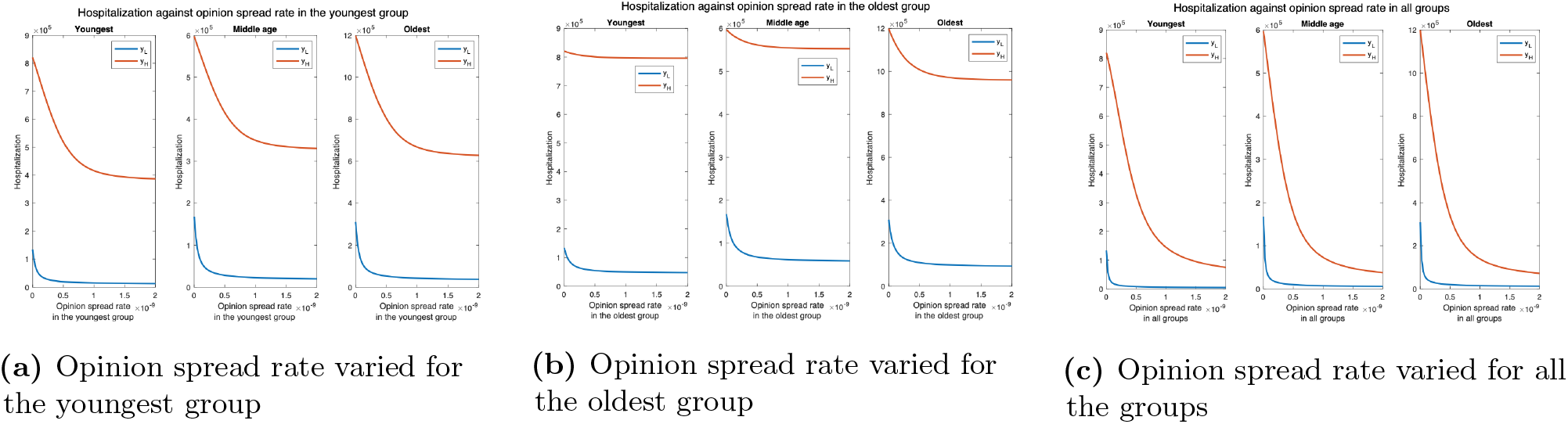
Hospitalization count for various interaction rates of the youngest group. *y_L_* denotes the default interaction rates of the youngest group while *y_H_* represent when the youngest group interaction rates are tripled (other groups interaction rates remain the same).

**Fig 19.**
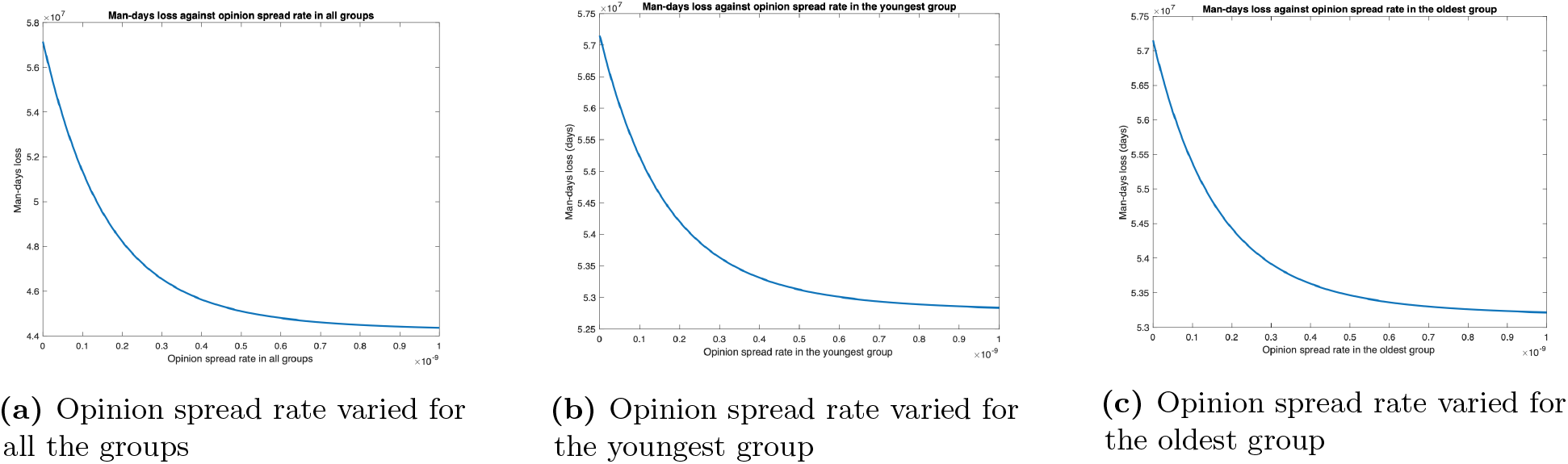
Impact of long COVID when vaccines are administered.

**Fig 20.**
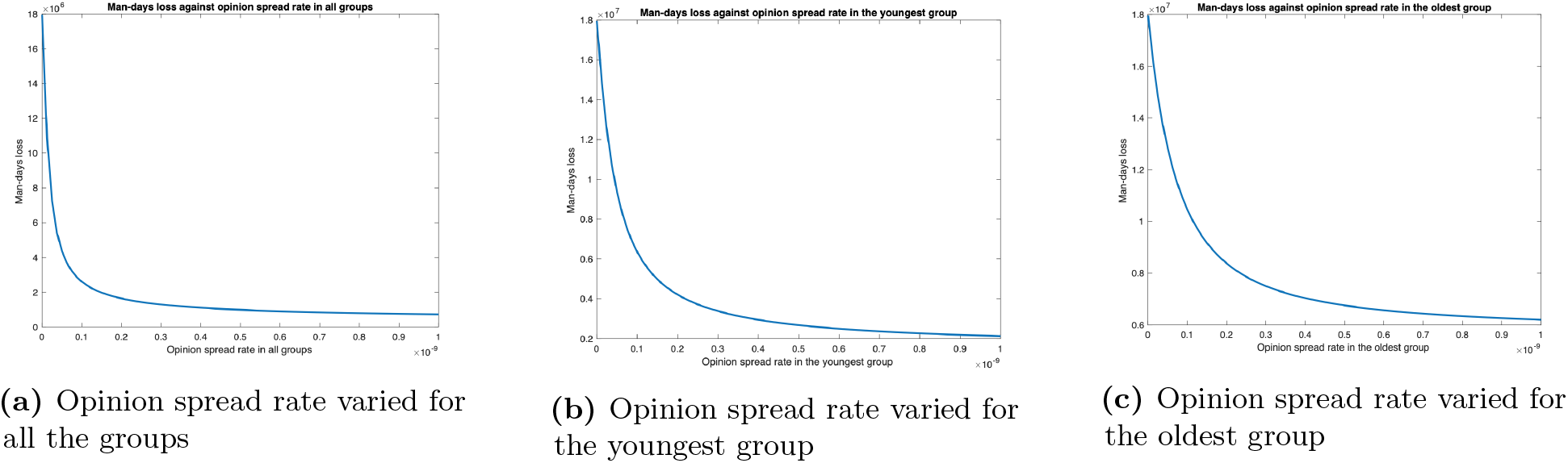
Impact of long COVID for the joint implementation of vaccines and behavioral practices.

